# Multi-omic Profiling of Recurrence Risk Across Breast Cancer Subtypes

**DOI:** 10.64898/2026.07.10.26357777

**Authors:** A. Eden Cruikshank, Pooja Chandra, Christopher I. Li, Gavin Ha

## Abstract

Recurrence risk greatly varies across intrinsic subtypes in breast cancer, yet the molecular and immune programs within primary tumors that go on to develop recurrence remains poorly understood. We performed multi-omic analysis of 340 breast cancer tumors across Basal-like, Luminal A, and Luminal B subtypes to identify tumor-intrinsic and microenvironmental features associated with recurrence. Within each intrinsic subtype, we compared recurrent and non-recurrent tumors across RNA, copy-number, and pathway-level mutational features. Basal-like tumors in patients who developed recurrence were characterized by reduced lymphocytes and pro-inflammatory M1 macrophages, enrichment of TGF-β/EMT activity, copy number gains within 5p/7p/7q, 4q losses, and increased pathway tumor mutational burden (pTMB) in growth-factor, inflammatory, and motility-associated signaling pathways, each of which was associated with increased recurrence risk. In Luminal A tumors, recurrent cases showed higher lymphocytes and pro-inflammatory M1 macrophages, enrichment of metabolic, stress-response, and stemness/plasticity associated pathways, and higher pTMB in growth-factor, inflammatory, motility-associated, DNA repair and apoptosis signaling pathways all associated with recurrence risk. Among Luminal B tumors, recurrent cases were enriched for proliferation, genomic instability, DNA repair, and stress-response pathways, showed a prominent 1q copy number amplification, and exhibited increased pTMB in Hedgehog signaling which increased recurrence risk. Subtype-specific prediction models were developed to generate recurrence-risk scores and validated using an external cohort (METABRIC; 1,170 total cases). The performance of our recurrence risk scores in METABRIC were associated with recurrence free survival (RFS) across subtypes (Basal: HR=1.27, 95% CI [1.07-1.50], p=0.006, Luminal A: HR=1.18, 95% CI [1.06-1.31], p=0.002, Luminal B: HR=1.41, 95% CI [1.03-1.93], p=0.03). Together, these findings demonstrate that primary tumors in patients who develop recurrence harbor distinct subtype-specific biological programs detectable at diagnosis and support a subtype-informed multi-omic modeling as a framework for recurrence-risk stratification.

## INTRODUCTION

Breast cancer remains a major clinical problem despite substantial improvements in detection and treatment. In the United States, an estimated 321,910 women will be diagnosed with invasive breast cancer and 42,140 women will die from breast cancer in 2026.^1^ Recurrence risk is not uniform across breast cancer but varies by intrinsic subtype. PAM50-based studies have shown that Basal-like and Luminal B tumors are associated with higher recurrence risk than Luminal A tumors.^2^ Additionally, timing differs by subtype: basal-like tumors most frequently recur within five years of diagnosis; while for Luminal A and Luminal B tumors the recurrence rate holds relatively constant over time, but is higher for Luminal B than Luminal A tumors, resulting in recurrences that can occur decades after initial diagnosis.^3^ These subtype-specific recurrence patterns underscore the need for recurrence-risk approaches that account for intrinsic tumor biology rather than treating breast cancer as a single clinical entity.

Recurrence biology is unlikely to be captured by a single molecular feature or clinical variable. Instead, metastatic relapse reflects coordinated processes with the tumor and its microenvironment, including tumor-cell plasticity, proliferation, genomic instability, metabolic adaptation, immune surveillance, and immune evasion. The tumor microenvironment contributes to multiple stages of cancer progression, from invasion and dissemination to metastatic outgrowth, while epithelial–mesenchymal plasticity can promote both metastatic competency and immune evasion. ^4,5^ In breast cancer specifically, immune context is prognostic and interacts with tumor-cell phenotypes such as EMT and proliferation, and previous studies have identified prognostic breast cancer groups characterized by differences in cell-cycle and immune-related pathways.^6–9^ These findings support a framework in which recurrence risk is shaped by both intrinsic tumor programs and the immune microenvironment, motivating subtype-stratified analyses that integrate molecular, genomic, and immune features.

Existing recurrence-risk models and multigene assays have improved breast cancer risk stratification and treatment decision-making, but they may not fully capture the subtype-specific and multi-dimensional biology underlying recurrence. Established approaches primarily rely on clinical variables and/or tumor gene-expression signatures, and their performance may vary across clinically and molecularly defined subgroups.^10–15^ In particular, prior work has suggested that additional data types, including DNA copy-number alterations and mutation status, may be needed to build more robust prognostic models.^16–19^ More recent studies further emphasize that breast cancer prognosis and treatment response reflect integrated genomic, transcriptomic, immune, and tumor-microenvironmental features rather than a single molecular layer alone. ^20–24^ These observations support the need for recurrence-risk models that are developed within intrinsic subtype and incorporate multiple feature classes, including RNA expression, copy-number alterations, pathway-level mutation burden, immune microenvironment features, and clinical variables.

Here, we performed a subtype-stratified analysis of recurrence-associated biology in Basal-like, Luminal A, and Luminal B breast cancers defined using PAM50 intrinsic subtype classification.^25^ We first characterized tumor-intrinsic and immune microenvironmental features associated with recurrence, including RNA pathway activity, copy-number alterations, pathway-level mutation burden, and immune-cell microenvironmental features. We then developed subtype-specific recurrence-risk scores using integrated RNA, copy-number, mutation-derived, immune, and clinical features, in two separate approaches: a direct multi-omic integration and stacked ensemble modeling. Model performance was assessed internally using iterative 5-fold cross-validation and externally with METABRIC, a large breast cancer resource with integrated genomic and transcriptomic data and long-term clinical follow-up, as a validation cohort.^19,26^

## RESULTS

### Multi-omic profiling of primary tumors reveals subtype-specific recurrence-associated features

We analyzed 340 primary treatment-naïve breast tumors collected at the time of diagnosis with available recurrence status and PAM50 subtype assignments. Samples were distributed across Basal-like (n=143), Luminal A (n=97), and Luminal B (n=100) subtypes. Recurrent and non-recurrent cases were represented at relatively similar proportions for Basal-like (71 vs 72, respectively), Luminal A (36 vs 61), and Luminal B (52 vs 48). (**Fig. 1A**) Within each subtype, recurrence groups were broadly balanced across key clinicopathologic and demographic variables, through matching recurrent to non-recurrent patients on age, stage, primary cancer-directed treatment, and race/ethnicity (**Fig 1B; Supplementary Table 1**). We then performed subtype-stratified multi-omic comparisons between recurrent and non-recurrent tumors. This framework allowed us to identify both shared and subtype-specific molecular features associated with recurrence, which were then incorporated into subtype-specific models to predict risk of recurrence.

**Figure 1.**
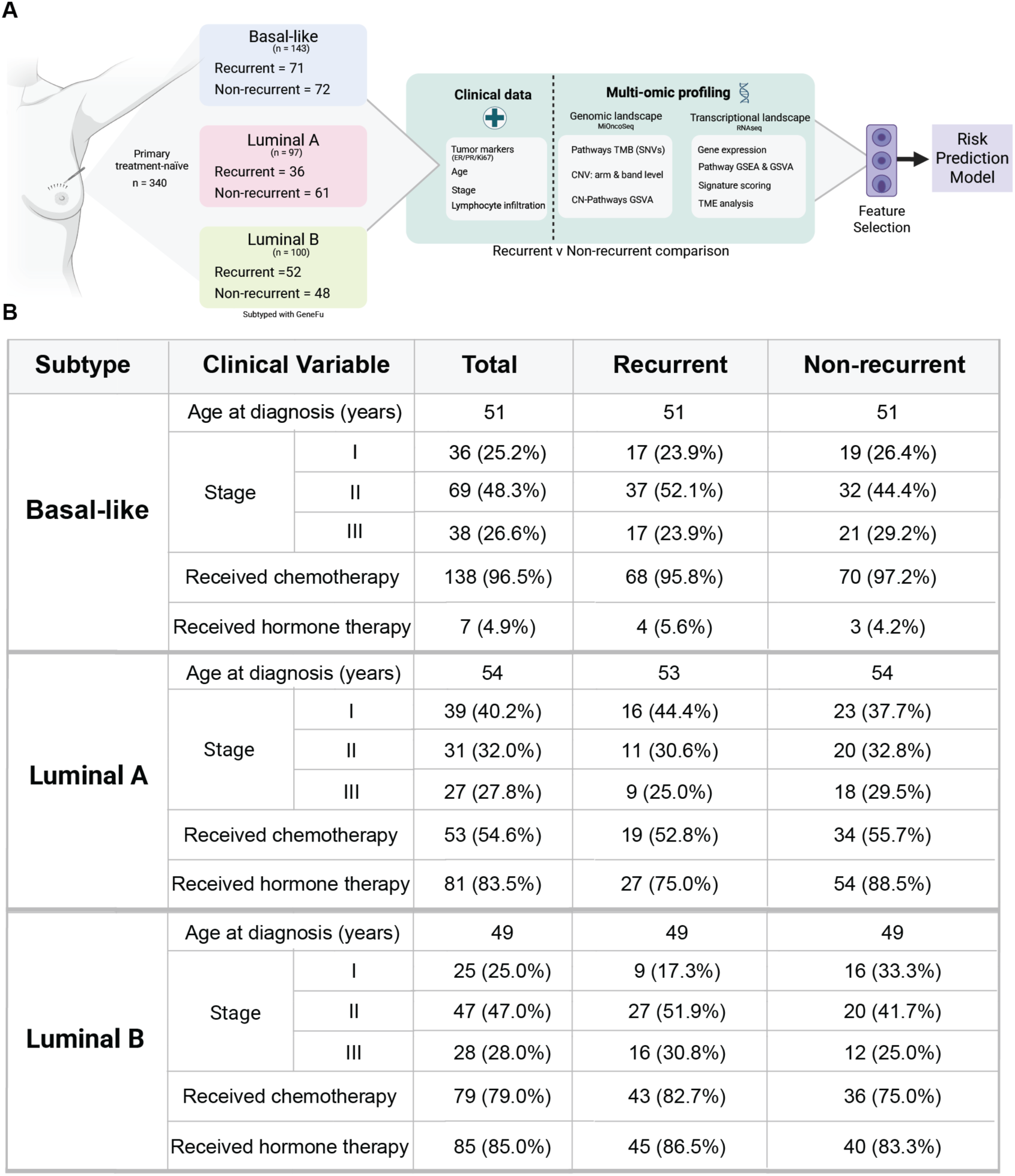
Study design and cohort characteristics. (a) Schematic overview of study design. A total of 340 primary treatment-naïve breast tumors were subtyped using GeneFu and stratified into Basal-like (n=143; recurrent=71, non-recurrent=72), Luminal A (n=97; recurrent=36, non-recurrent=61), and Luminal B (n=100; recurrent=52, non-recurrent=48) subtypes. Within each subtype, recurrent and non-recurrent tumors were profiled across clinical, genomic, and transcriptional features and compared to identify recurrence-associated biology, which was subsequently used to build subtype-specific recurrence-risk prediction models. (b) Clinicopathologic characteristics of the study cohort stratified by PAM50 subtype and recurrence status. Within each subtype, recurrent and non-recurrent groups were broadly balanced across age at diagnosis, disease stage, chemotherapy treatment, and hormone therapy treatment. Values represent counts with percentages in parentheses; age at diagnosis is reported as mean years.

### Recurrent tumors have intrinsic plasticity and markers of metastatic competency

To identify intrinsic molecular features associated with recurrence, we compared RNA pathway activity, copy-number alterations, and pathway mutational burden between recurrent and non-recurrent tumors within each PAM50 subtype (**Methods**). Transcriptional programs that were associated with recurrence were organized into three major groups across the three PAM50 subtypes: Luminal A-dominant pathways, Luminal B-dominant pathways with partial concordance in Basal-like tumors, and shared pathways across subtypes (**Fig. 2A**, **Supplementary Table 2**). The shared recurrence pathways captured broad cellular programs consistent with intrinsic plasticity and metastatic competency, including epithelial–mesenchymal transition (EMT), stromal remodeling, invasion, metabolic adaptation, and stress tolerance. Basal-like tumors which later recurred showed significance enrichment of independent pan-cancer EMT, epithelial-mesenchymal plasticity (EMP), and TGF-β-induced EMT signatures which were also associated with increased recurrence risk over time (**Fig. 2B, Supplementary Table 2**).^27–29^ Pathways enriched in Luminal B were characterized by enrichment of proliferative, growth, DNA repair (mismatch and nucleotide excision repair), RNA-processing, and stress-response programs. Luminal B tumors associated with recurrence had increased CLINSARC scores, a mesenchymal-tumor-derived sarcoma signature associated with chromosomal instability and metastatic risk (**Fig. 2C**, **Supplementary Table 2**).^30^ Pathways enriched in Luminal A showed a more heterogeneous recurrence-associated intrinsic profile, showing evidence of metabolic and stemness-associated plasticity and stromal-remodeling biology. Wnt-driven EMT and epithelial–mesenchymal plasticity signatures were directionally increased but less pronounced after adjustment for both hormone and chemotherapy treatment **(Fig. 2D, Supplementary Table 2)**.^28,31^

**Figure 2.**
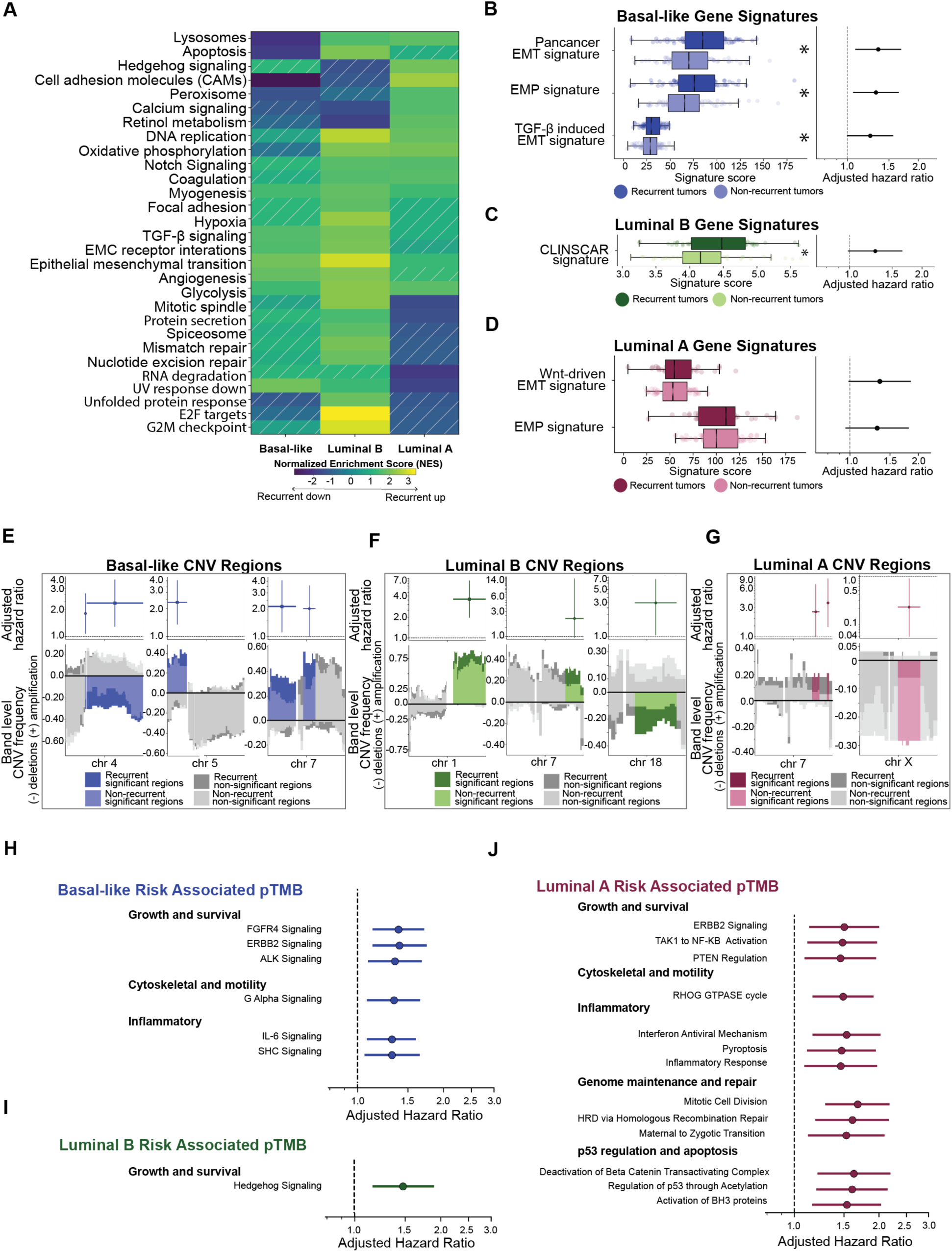
Recurrence-associated intrinsic transcriptional, copy-number, and mutational features across breast cancer subtypes. (a) Heatmap of recurrence-associated intrinsic pathway GSEA across Basal-like, Luminal A, and Luminal B subtypes. Color reflects direction of enrichment in recurrent tumors (blue = recurrent down, yellow/green = recurrent up). Cross-hatching indicates the pathway was not significant for in that subtype. (b–d) Recurrence-associated gene signatures for Basal-like (b), Luminal B (c), and Luminal A (d) subtypes. Box plots show signature score distributions in recurrent and non-recurrent tumors with corresponding adjusted hazard ratios for recurrence-free survival. Asterisks indicate statistical significance using Mann–Whitney U (MWU), p < 0.05. (**e–g)** Recurrence-associated copy-number regions for Basal-like (e), Luminal B (f), and Luminal A (g) subtypes. Bar plots show band-level CNV frequency across key chromosomal regions, with positive values indicating amplification and negative values indicating deletion. Colored bars indicate statistically significant regions by Cox regression; gray bars indicate non-significant regions. Adjusted hazard ratios for recurrence-free survival are shown for significant regions. (h–j) Recurrence-associated pathway-level tumor mutational burden (pTMB) for Basal-like (h), Luminal B (i), and Luminal A (j) subtypes. Forest plots show adjusted hazard ratios for recurrence-free survival across significantly enriched pathways grouped by biological category.

Next, we assessed copy number alterations associated with disease recurrence. We observed the enrichment of 5p and 7p/7q gains together with 4q losses which increased the risk of recurrence over time in patients with Basal-like tumors (**Fig. 2E**, **Supplementary Table 3**). These copy number gains contain genes involved in TGF-β/EMT regulation, metastasis, motility, cell invasion, and poor-prognosis (e.g. *RICTOR*, *SKP2*, *EGFR*, *RAC1*, *TWIST1*, and *ITGB8*), and are consistent with the transcriptionally altered programs we observed (**Supplementary Figure 1A**, **Supplementary Table 2)**.^32–39^ In Luminal B tumors, 1q amplification was associated with increased recurrence risk (**Fig. 2F**, **Supplementary Table 3**). This region contains KIF14 and RAB25, which we observed to have increased expression in Luminal B tumors from patients who recurred (**Supplementary Figure 1B**, **Supplementary Table 2**). ^40,41^ Additionally, we found a lower frequency gain at 7q36 containing *EZH2*, which was consistent with the upregulation of E2F target genes observed in Luminal B tumors from patients who recurred.^42,43^ Lower frequency recurrent-associated losses were also observed at 18q21 containing SMAD2/SMAD4, central mediators of TGF-β–driven EMT and metastatic progression.^44^ Luminal A tumors had more limited copy number alterations, and were driven primarily by smaller segmental events rather than broad arm-level aneuploidy. However, we did see amplifications at 7q36.1, containing *EZH2,* that increased the risk of recurrence over time, while Xq21–24 deletions were associated with reduced recurrence risk (**Fig. 2G, Supplementary Table 3, Supplementary Figure 1C**, **Supplementary Table 2**). ^42^

We assessed mutational burden of genes within molecular pathways (pTMB) to identify the impact of SNVs on the risk of recurrence. Basal-like tumors from patients who experienced recurrence showed increased pTMB across RTK–mTOR-axis, IL6, SHC, and Gα12/13 signaling pathways (**Fig. 2H**; **Supplementary Table 4**), implicating effects on growth-factor/survival, inflammation, and cytoskeletal/motility-associated signaling modules.^45–53^ Luminal B tumors only had an increase in pTMB in the Hedgehog pathway associated with risk of recurrence (**Fig. 2I**). Finally, pTMB in Luminal A tumors converged on several biological pathways, including RTK-mTOR axis and NF-KB signaling, genome maintenance and repair, p53 regulation and apoptosis, cytoskeletal/motility, and inflammatory stress signaling (**Fig. 2J**; **Supplementary Table 4**).^54–56^

Overall, the subtypes had genomic and transcriptional alterations that uniquely associated with disease recurrence, spanning pathways involved in mesenchymal transition and plasticity (Basal), genomic instability and stress-response (Luminal B), and metabolic adaptation, stemness-associated plasticity and inflammatory activation (Luminal A).

### Intrinsic subtype shapes the spectrum of immune microenvironments associated with recurrence

Next, we interrogated the immune microenvironment in the treatment-naïve tumors across subtypes to identify recurrence-associated immune biology. We first evaluated immune scores across recurrence groups and survival outcomes and identified the immune pathways that were associated with developing recurrence for each subtype (**Methods**).^57^ In Basal-like tumors, immune programs were less enriched in patients who recurred, and overall lower immune scores were associated with increased recurrence risk (**Supplementary Fig. 2A-C, Supplementary Table 5**). By contrast, Luminal A tumors showed broad enrichment of immune pathways in patients who recurred and exhibited higher immune scores that were associated with increased recurrence risk (**Fig. 3A**, **Supplementary Table. 2**). Interestingly, Luminal B tumors showed a mixed pattern for recurrent cases, with partial enrichment of inflammatory and immune-regulatory programs that were also observed in Luminal A, and downregulation of pathways such as JAK/STAT signaling and cytokine receptor interactions. Together, the spectrum of immune responses associated with recurrence is subtype-specific spanning broad immune activation (Luminal A), relative loss of immune activation (Basal), and a mixed inflammatory-regulatory state (Luminal B).

**Figure 3.**
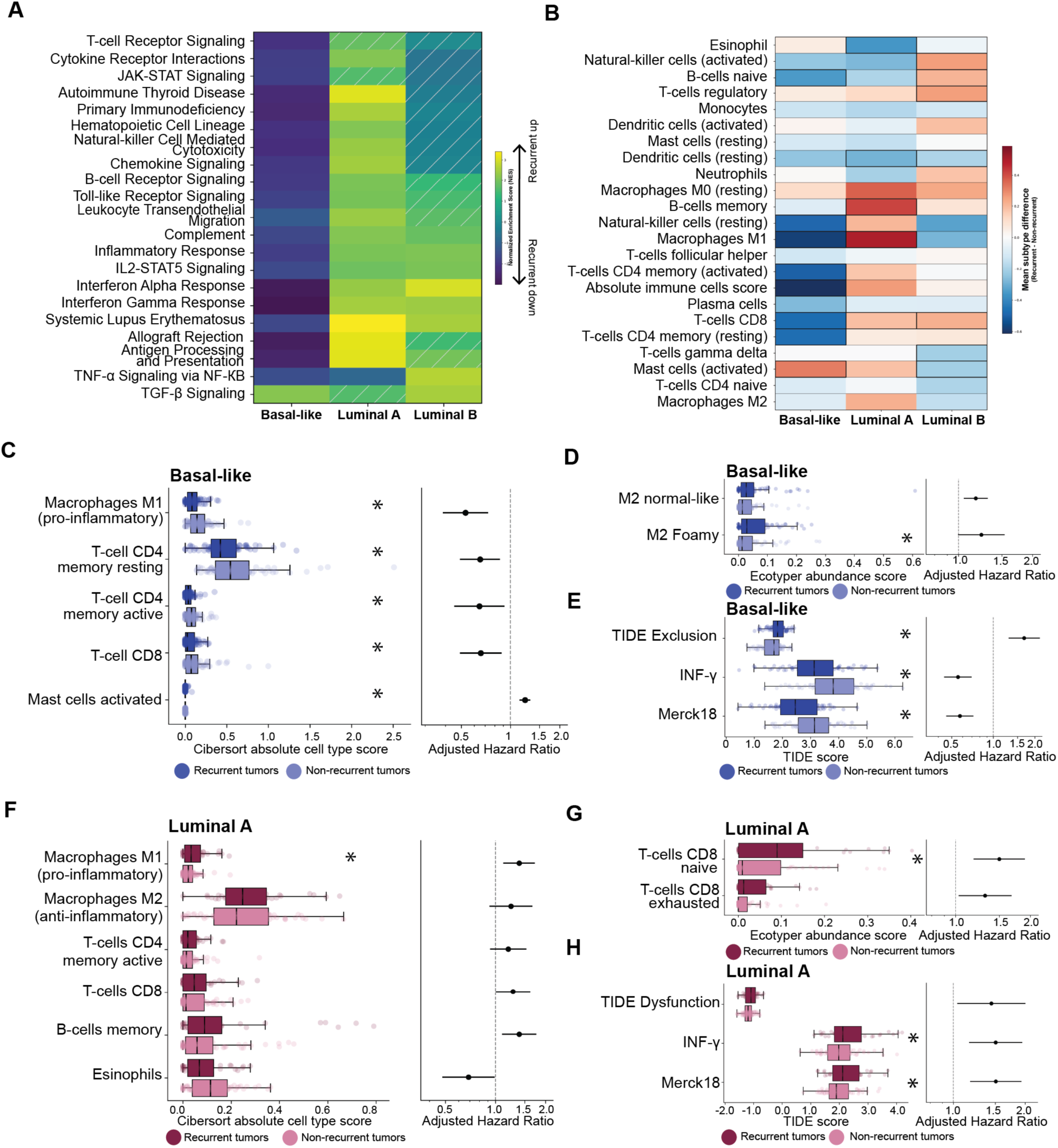
Recurrence-associated immune microenvironment features across breast cancer subtypes. (a) Heatmap of recurrence-associated immune pathway GSEA across Basal-like, Luminal A, and Luminal B subtypes. Color reflects direction of enrichment in recurrent tumors (yellow/green = recurrent up, blue/purple = recurrent down). Cross-hatching indicates the pathway was not significant for in that subtype. (b) Heatmap of mean zscore difference in CIBERSORT absolute immune cell type scores between recurrent and non-recurrent tumors across subtypes. Warm colors indicate higher abundance in recurrent tumors; cool colors indicate higher abundance in non-recurrent tumors. Outlined boxes indicate statistical significance by Cox regression. (c) Basal-like recurrence-associated CIBERSORT absolute scores. Box plots show immune cell type abundance in recurrent (dark blue) and non-recurrent (light blue) tumors with corresponding adjusted hazard ratios for recurrence-free survival. Asterisks indicate statistical significance by MWU. (d) Basal-like recurrence-associated EcoTyper macrophage cell state abundance scores, displayed as in (c). (e) Basal-like recurrence-associated TIDE immune evasion scores, INF-g scores and MERCK18 T-cell inflamed scores displayed as in (c). (f) Luminal A recurrence-associated CIBERSORT absolute scores. Box plots show immune cell type abundance in recurrent (dark pink) and non-recurrent (light pink) tumors with corresponding adjusted hazard ratios for recurrence-free survival. Asterisk indicates statistical significance by MWU. (g) Luminal A recurrence-associated EcoTyper T cell state abundance scores, displayed as in (f). (h) Luminal A recurrence-associated TIDE T-cell dysfunction scores, INF-g scores and MERCK18 T-cell inflamed scores displayed as in (f).

The tumor microenvironment showed divergent recurrence-associated features between Basal-like and Luminal A tumors specifically in immune cell compositions, immune cell states, and T cell activity (**Methods**). In Basal-like tumors, there was a broad immune-inflamed phenotype with higher T cell and M1 macrophage cell fractions that were associated with reduced recurrence risk, while activated mast cells increased the risk of recurrence (**Fig. 3B-C**, **Supplementary Table 5**). Further analysis of the tumor microenvironment revealed an M2 macrophage ecosystem with increased normal-like and foamy cells associated with recurrence risk, suggesting their immunoregulatory support of immune evasion and suppression (**Fig. 3D, Supplementary Table 6**).^58–60^ Furthermore, in cases that developed recurrence, Basal-like tumors were enriched for a carcinoma state linked to wound-healing, and squamous / Basal-like, programs (**Supplementary Figure 2F, Supplementary Table 6**).^61^ Analysis of immune evasion in Basal-like tumors revealed higher T cell exclusion was associated with increased recurrence risk, while T-cell inflamed scores (Merck18) and interferon-gamma signatures were associated with reduced risk (**Fig. 3E, Supplementary Table 7**).

Luminal A tumors showed a distinct recurrence-associated immune pattern (**Fig. 3B & Fig. 3F; Supplementary Table 2**). Recurrent associated tumors demonstrated broad immune recruitment of total macrophages (M0, M1, and M2 macrophage states), although only enrichment of M1 macrophages remained independently associated with recurrence (**Fig. 3F, Supplementary Figure 2D Supplementary Table 5**). Notably, M2 macrophages were more abundant overall in Luminal A than in either Basal-like or Luminal B tumors, suggesting that recurrence-associated immune recruitment in Luminal A occurs within a M2-rich baseline (**Supplementary Table 6, Supplementary Figure 2E**). Increased levels of CD4 memory active cells, memory B cells and CD8 T cells, specifically naïve and exhausted states, were associated with recurrence risk suggesting this immune-active state may be functionally constrained, whereas increased eosinophils were protective (**Fig. 3F-G, Supplementary Table 5-6**). Analysis of immune dysfunction showed higher T cell dysfunction, inflammation, and interferon-gamma signatures were associated with increased recurrence risk (**Fig. 3H, Supplementary Table 7**). Together, these findings suggest that immune recruitment in Luminal A tumors may not translate into effective anti-tumor immunity in patients who recur.^62^

Collectively, these analyses indicate that that immune activation was associated with better outcomes and reduced recurrence risk in Basal-like tumors, while immune recruitment, macrophage enrichment, and increased naïve and exhausted T cell dysfunction were associated with developing recurrence in Luminal A breast cancer.

### Integrated multi-omic recurrence-risk scores stratify outcomes across breast cancer subtypes

To determine whether recurrence-associated molecular and clinical features could be used to predict future recurrence, we developed multi-omic models to predict subtype-specific recurrence risk scores. For each subtype, we evaluated two modeling strategies: (1) all molecular and clinical features analyzed jointly in a single model (ALL model), and (2) features analyzed independently using separate models, with outputs combined into a single stacked meta-model (Ensemble model) (**Fig. 4A-B**, **Methods, Supplementary Table 8**). For Basal-like tumors, the ALL model showed stronger internal stability than the Ensemble model, producing a risk score that was significantly associated with recurrence-free survival (RFS), (HR=9.40, 95% CI [3.11-23.83], p<0.05 in 95/100 of cross-validation iterations) (**Fig. 4C**, **Supplementary Figure 3A, Supplementary Table 8**). Evaluating the features contributing to the model using SHAP revealed predominantly RNA-derived predictors, including RNA features linked to metastasis, EMT regulation, cell-cycle control, DNA-damage response, T-cell exhaustion, as well as copy-number pathways related to ion transport, metabolism, and immune regulation (**Fig. 4F, Supplementary Figure 3D**).^63–72^ For Luminal A tumors, both ALL and Ensemble models showed broadly similar internal performance, with risk scores associated with RFS (HR=3.91, 95% CI[1.35-11.65], p<0.05 in 35/100 iterations; HR=3.74, 95% CI[1.40-13.00], p<0.05 in 35/100 iteration, respectively) (**Fig. 4D**, **Supplementary Figure 3B, Supplementary Table 8**). Similarly, the recurrence risk in the model was primarily driven by RNA features in the ALL model, although select copy-number and mutations as well as age at diagnosis were also contributors. Many of the top features across datatype had prior links to luminal-to-basal plasticity, EMT, metastasis, endocrine or chemotherapy resistance, angiogenic remodeling, aggressive breast cancer biology and progression.^73–81^ (**Fig. 4G, Supplementary Figure 3E**). In Luminal B tumors, only the ALL model was associated with risk of recurrence (HR=5.41, 95% CI [1.00-15.93], p<0.05 in 41/100 iterations) (**Fig. 4E**, **Supplementary Figure 3C, Supplementary Table 8**). Again, RNA represented the bulk of contributing features in the ALL model. Several features had prior links to metastatic progression, endocrine or tamoxifen resistance, and poor outcomes (**Fig. 4H**, **Supplementary Figure 3F**). ^82–87^

**Figure 4.**
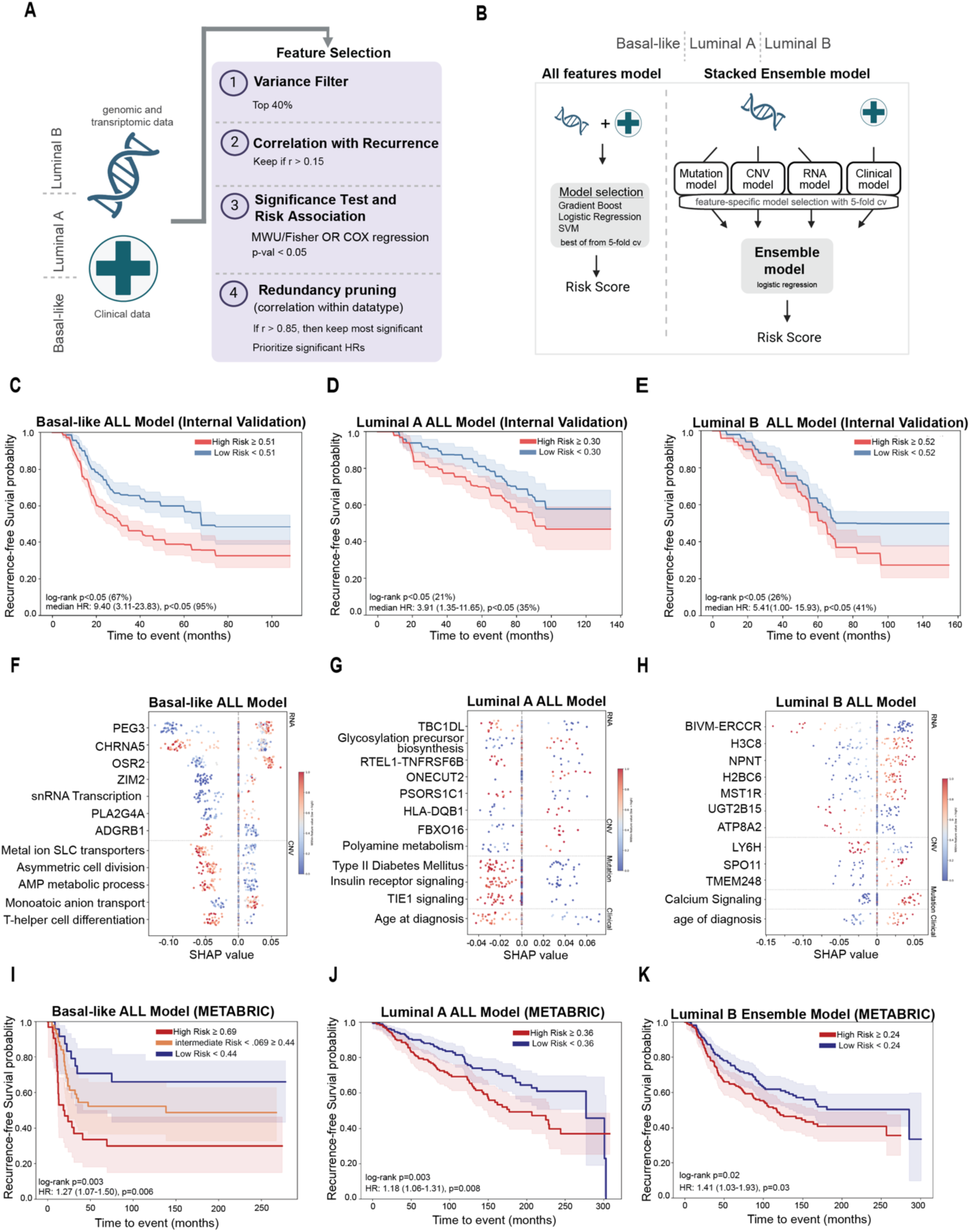
Development and validation of subtype-specific multi-omic recurrence-risk models. (a) Schematic of the four-step feature selection pipeline applied within each PAM50 subtype, filtering by variance, correlation with recurrence, significance and risk association, and redundancy pruning. (b) Schematic of the two modeling strategies evaluated per subtype: an ALL-features model jointly integrating genomic, transcriptomic, and clinical features, and a Stacked Ensemble model combining separate mutation, CNV, RNA, and clinical models via L2 logistic regression. (c-e) Internal validation Kaplan-Meier curves for recurrence-free survival stratified by risk score in Basal-like (c), Luminal A (d), and Luminal B (e) subtypes. Percent of log-rank p-values <0.05, median hazard ratios with 95% confidence intervals, and median high risk / low risk cut offs are shown. Shaded regions represent 95% confidence intervals. (f–h) SHAP feature importance plots for Basal-like (f), Luminal A (g), and Luminal B (h) recurrence-risk models. Points represent individual samples separated by data modality (RNA, CNV, mutation, clinical), with SHAP values indicating direction and magnitude of contribution to the risk score. (i–k) External validation Kaplan-Meier curves in the METABRIC cohort for Basal-like (i), Luminal A (j), and Luminal B (k) subtypes. The Basal-like model stratified patients into three risk groups; Luminal A and Luminal B into two risk groups. Log-rank p-values and hazard ratios based on continuous (Luminal A & Basal) or binarized (Luminal B) recurrence risk scores with 95% confidence intervals are shown.

To validate each subtype-specific recurrence risk model, we applied the independently trained models to the METABRIC breast cancer cohort. For the Basal-like subtype (n=147), the ALL recurrence-risk score significantly stratified patients into low-(25^th^ percentile), intermediate-, and high-risk (75^th^ percentile) groups (multi-group log-rank test p=0.003) and were associated with RFS (HR=1.27, 95% CI [1.07-1.50], p=0.006), (**Fig. 4I, Supplementary Figure 3G**). For the Luminal A subtype (n=460), the recurrence-risk score significantly stratified patients into low-, and high-risk groups split by median, and were significantly associated with RFS in both the ALL and Ensemble models (log-rank tests p=0.003 & p=0.007; HR=1.18, 95% CI [1.06-1.31], p=0.002; HR=2.37, 95% CI [1.36-4.12], p=0.002, respectively), (**Fig. 4J**, **Supplementary Figure 3H**). For the Luminal B subtype (n=563), only the Ensemble model using binarized risk scores (median) were associated with RFS (HR=1.41, 95% CI [1.03-1.93], p=0.03) and stratified recurrence-risk scores into low-, and high-risk patient groups split by median (log-rank test p=0.02), while the ALL model was less effective at stratifying outcomes (HR=1.31, 95% CI [1.00-1.71], p=0.05; log-rank test p=0.04) (**Fig**. **4K**, **Supplementary Figure 3I**). This suggests that recurrence risk in Luminal B tumors involves a more heterogenous set of complementary signals from each separate feature type. Altogether, the prediction of this risk score required distinct multi-omic modeling strategies because each subtype was characterized by unique combinations of molecular and clinical feature modalities that contributed to disease recurrence.

## DISCUSSION

In this study, we used a multi-omic framework to characterize subtype-specific recurrence-associated biology and build subtype-specific recurrence-risk models that significantly stratify outcomes in an independent validation cohort. Our findings demonstrate that recurrence is not driven by a single conserved program, but by distinct subtype-specific combinations of tumor-intrinsic plasticity, immune microenvironment, and clinical risk. In patients with Basal-like breast cancer who later recurred, tumors showed reduced immune signals and increased EMT/ TGF-β activity. Whereas Luminal A recurrence was characterized by increased immune signals, metabolic pathway expression, and stemness-associated alterations. Luminal B recurrent cases showed higher proliferation, genomic instability and stress adaptation features. Due to their subtype-specific biology, capturing risk required subtype-specific modeling strategies tailored to the distinct recurrence architecture of each subtype.

Basal-like recurrence was characterized by a transcriptional and genomic program of intrinsic metastatic competence, together with T-cell exclusion features and an immunosuppressive, wound-healing myeloid environment.^5,88–91^ These tumors exhibited a strong TGF-β-driven EMT signatures, with co-enrichment of extracellular matrix remodeling, angiogenesis, and glycolysis pathways. This suggests these tumors may already occupy a state of metastatic competence at diagnosis as EMT and epithelial–mesenchymal plasticity are linked to cell motility, invasion, stem-like behavior, dissemination, and metastasis. ^92,93^ Genomic findings reinforced this biology: recurrence-associated gains involving RICTOR/SKP2 (5p) and the 7p region (EGFR, RAC1, TWIST1, ITGB8) implicate TGF-β/EMT-associated programs, with RICTOR/mTORC2 promoting Rac1-dependent metastasis, SKP2 linked to EMT and therapy resistance, and high EGFR copy number predicting poor outcome in TNBC.^32–39,47^ Further, we saw recurrence-associated pTMB in RTK–mTOR, IL6/SHC, and Gα12/13 signaling alterations, implicating growth-factor signaling, JAK–STAT3 inflammatory activity, and Rho/ROCK/JNK cytoskeletal rewiring all of which have established roles in proliferation, invasion, immune suppression, and metastatic progression. ^45–53,94,95^ This intrinsic metastatic program appeared to extend beyond the tumor cell to reshape the immune microenvironment, as TGF-β has been shown to restrict CD8+ T-cell infiltration and polarize macrophages toward suppressive populations. ^5,88–90^ We observed evidence of both in our tumor microenvironment analysis. For example, Basal-like recurrence was characterized by impaired cytotoxic T-cell access, as shown by increased T-cell exclusion scores.^62,96^ We also observed an enrichment of activated mast cells and M2 normal-like and foamy macrophage states, which have been linked to angiogenesis, stromal remodeling, immunosuppression, and poor outcome.^58–60,97–105^ Additionally, we saw an enrichment of the wound-healing, squamous/basal-like associated carcinoma state; a phenotype associated with immune suppression and tumor progression.^106–109^ This contrasts with Basal-like tumors that did not recur which were enriched for immune-active features including CD8+ and CD4+ T-cell subsets, B cells, and M1 macrophages, consistent with prior literature linking higher TIL abundance and cytolytic activity to improved OS and DFS in TNBC. ^110–115^ These non-recurrent cases also exhibited lower TGF-β activity which is associated with increased lymphocyte activity and more favorable outcomes, as shown in our data and in previous literature.^91^ Together, these transcriptional, copy-number, and pathway-level signals converge on a TGF-β-driven EMT program intertwined with myeloid-stromal remodeling and immune exclusion suggesting metastatic competence in recurrent Basal-like disease.

However, in Luminal A breast cancer, immune engagement appeared to mark increased recurrence risk rather than protection. In our cohort, we saw higher M1 macrophages, CD8 T cells, memory B cells, TIDE Dysfunction, Merck18, and IFN-G all associated with increased recurrence risk. Enrichment of both naive and exhausted CD8 T cells in recurrent tumors may suggest T cells are being recruited but failing to activate or become dysfunctional. This is consistent with literature showing high TILs are not clearly protective in luminal disease and may associate with worse OS, and with reports linking CD8 infiltration and exhaustion signatures to increased recurrence risk in HR-positive disease.^116–118^ Additionally, recurrent Luminal A tumors showed broad enrichment across macrophage states and higher overall M2 abundance than Basal-like or Luminal B tumors, suggesting immune recruitment occurs within a higher M2 baseline. TAM/M2-like macrophages have been shown to suppress CD8 T-cell function, promote angiogenesis, and drive extracellular matrix remodeling, pathways which were transcriptionally enriched in our Luminal A recurrent cases.^102,104,119–122^ Although M1 macrophages were the only macrophage state independently associated with recurrence in our data, this likely reflects inflammatory activation within a pro-tumor macrophage environment rather than productive anti-tumor immunity. Together, all of this suggests that immune features detected at diagnosis reflect an inflammatory microenvironment with immunosuppressive features rather than an anti-tumor response.

In contrast to the EMT driven program of recurrence Basal, Luminal A cases exhibited a weaker mesenchymal signature with enrichment in metabolic, stemness-associated signaling, repair, and inflammatory-stress pathways. This aligns with prior work showing that ER-positive breast cancer can enter dormancy marked by epithelial–mesenchymal plasticity which we saw was directionally increased in recurrent cases.^123^ Metabolic enrichment in oxidative phosphorylation, TCA cycle, glycolysis, and peroxisome pathways suggests metabolic flexibility allowing survival under therapeutic pressure, consistent with studies reporting oxidative phosphorylation dependence in metastatic, therapy-resistant ER-positive disease.^86,124–127^ Recurrence-associated RTK/mTOR, Hedgehog, IL6–JAK–STAT3, repair, and p53/apoptosis pTMB enrichment further supports a model of signaling flexibility, stem-like plasticity, and stress tolerance.^48,128–135^ Additionally, amplification of the region containing EZH2 (7q36.1) is consistent with literature linking this epigenetic regulator to poor outcomes in Luminal A disease.^42,136^ Together, these multi-omic signals of metabolic flexibility, adaptive signaling, and stemness leads to a baseline tumor state primed for long-term persistence, and recurrence in Luminal A tumors.

Luminal B tumors in patients which later recurred were primarily defined by a highly proliferative, genomically unstable, and stress-adapted biology, with immune signals playing a secondary and mixed role rather than defining the recurrence phenotype. Recurrent tumors showed enrichment of proliferative and growth (E2F targets, G2M checkpoint), DNA repair (mismatch and nucleotide excision repair), and stress-response programs (hypoxia and angiogenesis). Prior work linking E2F and G2M activity to aggressive and metastasis-associated biology in ER-positive breast cancer, and increased CINSARC scores being associated with Luminal B treatment resistance further support chromosomal instability as a defining feature.^43,90,137–140^ The copy-number landscape reinforced this state: dominant 1q amplification is consistent with prior studies linking 1q alterations to metastatic progression, with recurrence-associated regions containing KIF14, S100 genes, and RAB25, shown to promote EMT, while EZH2 amplification within 7q36 and SMAD2/SMAD4 loss within 18q implicate chromatin-mediated plasticity and TGF-β dysregulation as potential recurrence mechanisms.^40,43,44,87,136,141–147^ Notably, EZH2 lies downstream of the pRB–E2F pathway and is directly regulated by E2F, linking the genomic and transcriptional findings.^43^ The immune phenotype was mixed, consistent with prior work describing Luminal B immune contexture as an intermediate inflammatory/immunoregulatory state rather than a clearly protective anti-tumor response.^148,149^ Together, these findings suggest that recurrent Luminal B tumors acquire metastatic competence primarily through replicative stress, chromosomal instability, and plasticity.

We generated subtype-specific recurrence-risk scores that significantly stratified recurrence-free survival in an independent validation cohort, with each subtype requiring distinct multi-omic modeling strategies. Current clinically approved tests are largely limited to ER-positive disease, have no established utility in TNBC, and rely on single-platform RNA-based features that do not capture the multi-omic signatures associated with recurrence identified here, which includes copy-number and mutation-derived features.^10,13,150,151^ While, prior work from our group characterized intrinsic and immunologic phenotypes in Basal-like disease, and Prosigna/PAM50 provides molecular subtyping each with their own recurrence risk, neither addresses subtype-specific recurrence prediction within all three subtypes.^8,152–154^ In Basal-like tumors, the ALL model generalized best, driven by a strong transcriptomic signal with copy-number features providing complementary information. In Luminal A, both ALL and ensemble models validated externally, reflecting a distributed risk architecture spanning RNA, copy-number, mutation-derived, immune, and clinical features. In Luminal B, only the stacked model generalized to external validation, suggesting that mutation-derived, clinical, and copy-number signals better captured recurrence risk than a dominate RNA feature space. Importantly, across the subtypes, top model features had previously established links to breast cancer progression, metastasis, EMT/plasticity, angiogenesis, and treatment resistance. ^63–87^ Thus, even in Luminal B, where performance was limited, the selected features still highlighted biologically relevant processes rather than model noise. However, the weaker and mixed performance in Luminal B may reflect its clinical heterogeneity and studies with larger Luminal B cohorts that can be stratified by key clinical features may better resolve recurrence risk in the future. Together, these findings demonstrate that disease recurrence has distinct molecular underpinnings across subtypes, and that subtype-specific multi-omic integration captured recurrence risk beyond single-platform approaches.

Several limitations of this study warrant consideration. While the cohort in this study included detailed, curated recurrence and treatment data, the analysis was retrospective and observational, and there were confounding factors such as treatment history, clinical management, and follow-up that cannot be fully excluded. Although cohort balancing and adjustment for chemotherapy and endocrine therapy helped to account for potential treatment biases, therapies were administered after primary tumor collection which made it difficult to disentangle recurrence-free survival from treatment response. Subtype stratification also reduced discovery sample sizes, particularly for Luminal A tumors, limiting statistical power to detect weaker effects and treatment interactions. Finally, the recurrence-associated features and risk models reported here are prognostic rather than predictive, and functional and prospective validation will be needed to determine whether they directly contribute to tumor metastatic potential and disease aggressiveness.

Overall, these findings demonstrate that primary breast tumors which later recurred harbor subtype-specific biological programs detectable at diagnosis that are distinct, clinically meaningful, and not captured by existing single-platform or subtype-agnostic prognostic tools. These results make a strong case for moving beyond pan-breast recurrence models toward subtype-informed frameworks that integrate tumor-intrinsic, immune microenvironment, and clinical features, and lay the groundwork for prospective validation of subtype-specific recurrence biology as a foundation for recurrence risk stratification in breast cancer.

## METHODS

### Sex as a biological variable

Our study exclusively focused on female patients diagnosed with breast cancer because the disease primarily effects female patients (>99%).

### Statistics

Power calculations were performed separately for each subtype-specific discovery cohort based on the number of recurrent and non-recurrent tumors available for analysis. Using a one-sided test of AUC > 0.50 at α = 0.05, the Basal-like cohort had approximately 99.7% power to detect a discriminatory biomarker with AUC = 0.70 and 93.8% power to detect a biomarker with AUC = 0.65. The Luminal A cohort had approximately 96.0% and 80.0% power to detect biomarkers with AUC values of 0.70 and 0.65, respectively. The Luminal B cohort had approximately 97.8% and 84.1% power to detect biomarkers with AUC values of 0.70 and 0.65, respectively. For binary biomarkers with high specificity, AUC values of 0.70 and 0.65 correspond approximately to sensitivities of 40% and 30%, respectively.

Subtype specific statistical analysis was performed to evaluate associations between each variable and recurrence status or RFS. Continuous variables were compared between recurrent and non-recurrent cases using Mann Whitney U test. Binary categorical variables were evaluated with Fisher’s exact or chi-square tests. Cox proportional hazards regression was performed for all variables to assess associations with RFS, adjusting for age and treatment. Multiple test correction was performed for Cox proportional hazards regression and non-survival statistical comparisons with Benjamini-Hochberg false discovery rate (FDR) method. Statistical significance was defined as nominal P < 0.05 and FDR < 0.25. Thresholds used for model selection are described separately in the model development section of the methods.

### Study Approval

All study procedures were reviewed and approved by the Fred Hutchinson Cancer Center Institutional Review Board and were conducted in accordance with U.S. Common Rule guidelines. Informed consent was obtained from all living participants, and eligible deceased participants were enrolled through an IRB-approved waiver of consent, reducing survival bias by allowing tumor tissue collection and medical record review for deceased patients.

### Overall study design

To discover and externally validate subtype-specific recurrence-risk signatures, we analyzed three subtype-defined discovery cohorts from the Breast Cancer Risk Factors and Various Outcomes (BRAVO) breast cancer cohort and one independent external validation cohort, METABRIC. The discovery cohorts consisted of primary breast tumors stratified by PAM50 subtype into Basal-like, Luminal A, and Luminal B groups. PAM50 subtypes were derived using the Genefu R package. Recurrent and non-recurrent tumors were represented at relatively similar proportions in the Basal-like cohort (71 recurrent and 72 non-recurrent tumors), Luminal A cohort (36 recurrent and 61 non-recurrent tumors), and Luminal B cohort (52 recurrent and 48 non-recurrent tumors).

We used the BRAVO cohort for discovery because it is highly annotated and includes manually curated medical record data, allowing detailed ascertainment of recurrence outcomes, recurrence-free survival time, primary treatments, demographics, clinical characteristics, and tumor features. Details of this cohort have been described previously^8,155,156^ Briefly, women 20-69 years of age with newly diagnosed first primary invasive stage I-III breast cancer were identified through the Cancer Surveillance System, the NCI-funded population-based Surveillance, Epidemiology, and End Results cancer registry serving western Washington state.

External validation was performed in METABRIC, which was selected because it includes publicly available molecular data, PAM50 subtype annotations, clinical follow-up, and recurrence-associated survival outcomes. Final subtype-specific models were locked before application to METABRIC. No METABRIC samples were used for feature selection, model training, hyperparameter tuning, or model revision. Model-derived recurrence-risk scores were generated for METABRIC tumors within each corresponding subtype and evaluated for association with recurrence-free survival using Kaplan–Meier analysis and Cox proportional hazards regression.

### RNA and Targeted Panel library preparation and sequencing

Tumor total RNA was isolated from FFPE specimens using the AllPrep DNA/RNA/miRNA Kit (Qiagen). RNA sequencing was performed using an exome-capture transcriptome platform (Agilent SureSelect Exome V4) following standard protocols. DNA sequencing was performed using a targeted oncology panel, Roche Onco1500 v4; OncoPanelV4 or, Roche Onco1500 v2; OncoPanelV2. All sequencing was performed on FFPE-derived nucleic acids. All samples were sequenced on the Illumina HiSeq 2000 or HiSeq 2500 in paired-end mode.

### RNA-sequencing preprocessing, quality control, and differential expression analysis

Raw paired-end RNA-sequencing reads were assessed with FastQC, adapter- and quality-trimmed using Trim Galore, and filtered for ribosomal RNA using BBDuk. Filtered reads were quasi-mapped and quantified with Salmon against a GENCODE release 41 transcriptome index. Salmon files were imported using tximport, with transcript-to-gene mappings derived from the GENCODE v41 GTF and converted to gene-level count and abundance matrices. The TPM matrix was retained for downstream expression-based analyses and feature extraction.

RNA-seq quality control was performed using expression-based metrics, including total transcript counts, library complexity, and number of detected genes. Samples failing two or more QC metrics were excluded, resulting in removal of 23 RNA-seq samples from downstream analyses. Any remaining ribosomal genes were identified using an HGNC ribosomal gene list and removed from the count matrix prior to differential expression analysis as an additional measure to remove rRNA. RNA-seq quality control metrics, including sequencing (from Multiqc and BBDuk) and expression-based metrics (mentioned above), are provided in **Supplemental Table 9 and Supplemental Table 10**.

Differential expressions was performed separately within each PAM50 subtype using limma-voom. For each subtype, raw gene-level counts were normalized using edgeR TMM normalization, transformed using voom, and modeled with limma linear models followed by empirical Bayes moderation. The design matrix included recurrence status only, and the contrast was defined as recurrent versus non-recurrent disease. Genes were retained for testing if they had a total count of ≥50 and had ≥10 counts in at least 10% of samples. Differentially expressed genes were defined as those with nominal P < 0.05, absolute log2 fold change > 0.5, and base mean expression > 50. Gene-level Cox proportional hazards models were also fit using recurrence as the event and adjusting for age and treatment; genes were considered Cox-associated if P < 0.05. Genes were retained as candidate RNA model features if they met either the limma differential-expression criteria or the adjusted Cox regression criteria.

### Gene set enrichment analysis

Preranked GSEA was performed separately within each PAM50 subtype using recurrence-associated differential expression results. Genes were ranked by multiplying log2 fold change by the negative log10 nominal P value, generating a signed statistic in which positive values corresponded to genes increased in recurrent tumors and negative values corresponded to genes increased in non-recurrent tumors. Ranked gene lists were tested against KEGG and Hallmark gene-set collections. Pathways were considered enriched if nominal P < 0.05 and FDR q < 0.25.

For heatmap visualization, pathways were prioritized by significance, normalized enrichment score, and concordance across subtypes, then separated into intrinsic tumor-associated and immune-associated gene-set groups. Intrinsic and immune pathways were hierarchically clustered separately using subtype-specific normalized enrichment scores. Cross-hatched pathways did not meet the prespecified enrichment threshold but were retained to show directional concordance across subtypes; full NES, nominal P values, and FDR q values are provided in Supplementary Table 2.

### Gene set variation analysis

Sample-level pathway activity was quantified using GSVA on log2(TPM + 1) expression values for Hallmark, KEGG, and Reactome gene sets. Analyses were performed separately within each PAM50 subtype. GSVA scores were compared between recurrent and non-recurrent tumors, and pathway-level Cox proportional hazards models were fit using recurrence as the event, adjusting for age and treatment. Pathways were considered recurrence-associated if nominal P < 0.05 and FDR q < 0.25 in the group comparison or adjusted Cox analysis. Hazard ratios were used to indicate the direction and magnitude of recurrence-associated pathway activity. GSVA scores were incorporated as RNA pathway-level features in downstream recurrence-risk models.

### Molecular signature analysis

Published molecular signatures were scored from TPM-derived RNA expression values using either geometric mean log2TPM or median log2(TPM + 1) values across signature genes. Signatures included innate and adaptive enriched tumors, CLINSAR, and several Epithelial to Mesenchymal Transition / Plasticity signatures.^27–31,157–163^ Analyses were performed separately within each PAM50 subtype. Signature scores were compared between recurrent and non-recurrent tumors with Mann–Whitney U and were also evaluated using Cox proportional hazards models with recurrence as the event, adjusting for age and treatment. Signature scores were also binarized (median) and compared between recurrent and non-recurrent tumors with Fisher exact and Cox proportional hazards models with recurrence as the event, adjusting for age and treatment. Signatures were retained as candidate model features if either the group comparison or adjusted Cox regression met nominal P < 0.05.

### Tumor microenvironment analysis

Tumor microenvironment features were evaluated using RNA expression (log2(TPM + 1)) with CIBERSORT-derived immune cell fractions, EcoTyper cell-state abundances, and TIDE immune dysfunction/exclusion scores.^62,106,164^ CIBERSORT and EcoTyper Carcinoma Types were analyzed across PAM50 subtypes to assess overall subtype-associated immune composition differences. CIBERSORT, EcoTyper Carcinoma Types and EcoTyper Cell States, and TIDE scores were separately assessed within each subtype to evaluate the recurrence-associated tumor microenvironment. Within-subtype comparisons between recurrent and non-recurrent tumors were performed using the Mann–Whitney U test. Cox proportional hazards models were also fit using recurrence as the event, adjusting for age and treatment. Features were considered recurrence-associated if nominal P < 0.05 by either Mann–Whitney U testing or adjusted Cox regression. For heatmap visualization, CIBERSORT features were z-scored (within subtype) and differences in mean subtype-scaled feature abundance between recurrent and non-recurrent tumors were calculated. Hierarchical clustering was used to organize CIBERSORT features in heatmaps.

### Targeted panel alignment and preprocessing

Paired-end targeted-panel FASTQ files were processed using a Snakemake workflow. Reads were aligned to the reference genome using BWA-MEM, then converted to BAM format using SAMtools. BAM files were coordinate-sorted with SAMtools, and duplicate reads were marked using Picard MarkDuplicates. Base-quality score recalibration was performed using GATK BaseRecalibrator with known polymorphic sites specified in the workflow configuration, followed by ApplyBQSR to generate final recalibrated BAM files and indexes. Alignment quality metrics, duplicate metrics, and insert-size metrics were collected using Picard. Genomic quality-control metrics for targeted-panel sequencing including Picard HS, insert size and alignment metrics are in **Supplementary Table 11**.

### Targeted panel mutation calling and annotation

Somatic variants were called from targeted-panel tumor–normal BAM files using GATK Mutect2 and Strelka2 with matched-normal filtering. Variants were retained for downstream analysis only if they were identified by both callers. Consensus variants were annotated with ANNOVAR against hg38 using gene, population-frequency, cancer, clinical, and functional annotation databases, including RefGene, COSMIC, ClinVar, dbNSFP, InterVar, gnomAD, dbSNP, and 1000 Genomes resources. ANNOVAR annotation was performed with VCF input and nearest-gene annotation using a 5-kb window.

Consensus somatic variants were filtered to reduce germline contamination, low-confidence calls, and FFPE-associated artifacts. Variants were required to have AF_popmax <0.10, tumor and normal depth ≥10 reads, tumor variant allele fraction ≥0.02, normal variant allele fraction ≤0.10, tumor alternate depth ≥3 reads, and normal reference depth ≥8 reads. SOBDetector was applied to assess FFPE-associated strand-orientation artifacts, and variants predicted to represent FFPE artifacts were excluded before downstream mutation analyses.^165^ Because FFPE-associated artifacts are enriched for low-frequency C>T/G>A transitions, C>T and G>A variants were subjected to an additional VAF threshold. Consistent with prior FFPE filtering strategies, C>T/G>A variants were retained only if tumor VAF was ≥0.15 and they passed SOBDetector artifact filtering.^165,166^

### Pathway tumor mutational burden analysis

Pathway-level mutation features were generated from filtered consensus somatic SNVs. For each sample, pathway tumor mutational burden was calculated using an approach adapted from PathwayTMB, in which gene-level mutation density was calculated for each pathway gene and then summarized across the pathway:^167^

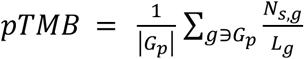

Where *s* is the sample, *p* is the pathway, *G_p_* is the set of genes in pathway *p*, *n_s,g_*, is the number of filtered SNVs in gene *g* for sample *s*, and *L_g_* is the coding length of gene *g*.

In parallel, binary pathway mutation features were generated to indicate whether any gene in a pathway was mutated in a given sample.^70^ Both continuous pathway TMB values and binary pathway mutation indicators were evaluated separately within each PAM50 subtype. Recurrence-associated differences were assessed using distributional testing between recurrent and non-recurrent tumors and Cox proportional hazard regression with recurrence as the event, adjusting for age and treatment. For overall significance analysis, pathways were pruned for redundancy by determining the overall overlap in genes between pathways and dropping pathways when overlap was greater than 0.75 of genes. Pathway mutation features were retained for downstream modeling if either the distributional comparison or adjusted Cox regression met nominal P < 0.05.

### Copy-number analysis

Copy-number profiles were generated from targeted-panel MiOncoSeq data using a modified CNVkit–ichorCNA pipeline. Because all samples were from female patients, chromosome X was retained and chromosome Y was excluded from downstream copy-number analyses. Gene-level copy-number values were normalized by sample ploidy and converted to copy-number deviation by subtracting 1, such that 0 represented copy-number neutral, positive values represented gains, and negative values represented losses. Gene-level gains and losses were defined using copy-number deviation thresholds of ≥0.3 and ≤−0.3, respectively. Cytoband-level copy-number features were generated by mapping genes to genomic coordinates and assigning genes to cytobands. For each sample and cytoband, binary amplification/deletion calls were calculated. Cytoband amplification or deletion was called when at least 60% of mapped genes in the band met the corresponding gain or loss threshold, and cytobands with fewer than three mapped genes were excluded. Arm-level and genome-level copy-number burden features were also calculated, including percent arm amplified, percent arm deleted, and percent genome altered. Arm-level events were binarized as present when the corresponding percent alteration was greater than 80%. To evaluate pathway-level copy-number patterns, GSVA was performed on the gene-level copy-number deviation matrix using variance-filtered pathway gene sets. This generated sample-level copy-number pathway activity scores, which were analyzed within each PAM50 subtype. Each copy number feature type was compared between recurrent and non-recurrent tumors within each subtype and were also evaluated using Cox proportional hazards regression with recurrence as the event, adjusting for age and treatment. Copy-number features were retained for downstream modeling if they met nominal P < 0.05 by either recurrence-group testing or adjusted Cox regression. Gene-level, cytoband-level, arm-level, genome-level, and CN-GSVA pathway-level copy-number features were included as candidate inputs for downstream recurrence-risk modeling.

### Risk Model Feature Selection

Feature selection was performed independently within each PAM50 subtype using the following sequential filtering: variance, correlation with recurrence, recurrence-association testing, and redundancy-pruning **(Fig.4A)**. Candidate RNA, copy-number, mutation, and molecular-signature features were first filtered using label-free variance criteria to remove low-variability predictors keeping only the top 40% of features. Features passing this step were then screened using a supervised feature–label correlation filter (r > 0.15), with recurrence encoded as a binary outcome. Remaining features were tested for recurrence association using two-sided Mann–Whitney U tests for continuous features and Fisher’s exact tests for binary features. In parallel, feature-wise Cox proportional hazards models were fit using recurrence as the event and adjusting for age and treatment covariates. Features were retained if they met nominal P < 0.05 by at least one recurrence-association test: Mann–Whitney U, Fisher’s exact test, or adjusted Cox regression. Selected features were then pruned for redundancy within molecular feature blocks using feature–feature correlation. When groups of highly correlated features exceeded the prespecified correlation threshold (0.90), a single representative feature was retained based on the strongest recurrence-associated evidence, prioritizing adjusted Cox regression results when available. This produced final non-redundant, subtype-specific feature matrices for downstream recurrence-risk modeling.

### Model training and model selection

Subtype-specific recurrence-risk classifiers were trained using the final post-feature-selection features. For each PAM50 subtype, recurrence status was encoded as a binary outcome, with recurrent tumors assigned to class 1 and non-recurrent tumors assigned to class 0. Candidate predictors included four feature blocks: RNA-derived features (including immune/TME features, molecular signature scores), copy-number features, mutation-derived features, and clinical covariates. Model-family selection was performed independently within each subtype and feature block using 5-fold cross-validation with ROC AUC as the optimization metric. Candidate model families were evaluated separately for RNA, copy-number, mutation, clinical, and concatenated ALL feature matrices. For each subtype/block combination, the model family with the highest mean cross-validated ROC AUC was selected and fixed for final training. The final locked model families for each stacked block (RNA, CNV, MUT & CLINICAL) as determined by 5-fold cv were Gradient Boosting for Basal RNA, mutation, and ALL features, ExtraTrees for Basal copy number, and L2-penalized logistic regression for Basal clinical features; LinearSVC for Luminal A RNA and clinical features, Random Forest for Luminal A copy-number and SVM for mutation features, and L2-penalized logistic regression for Luminal A ALL features; and Gradient Booting for Luminal B RNA features, Random Forest for Luminal B copy-number features, ElasticNet penalized logistric regression for Luminal B mutation features, and L1-penalized logistic regression for Luminal B clinical features and ALL features. Final hyperparameter tuning was then performed using GridSearchCV restricted to the selected model family, and the best model was refit using all available training samples within that subtype. Continuous molecular features were standardized using z-score scaling within the training data. Binary pathways mutations, clinical variables, discrete copy-number calls, and other binary/discrete features were passed through without scaling. Class imbalances were addressed during training using recurrent-class oversampling and class-weight tuning when supported by the model family. The two main integrated modeling strategies trained were the ALL model, trained as a single classifier using the full concatenated feature matrix across all retained molecular feature blocks, and a stacked ensemble model, trained using the feature block-level classifiers (as listed above). For the stacked model, each block-level classifier generated predicted probabilities of recurrence, and these probabilities were used as meta-features for either an L1 or L2-regularized logistic regression meta-model (Basal: L1, Luminal A: L1, Luminal B: L2). Final hyperparameter tuning of the meta model was performed using GridSearchCV. The primary model output (both ALL and stacked ensemble) was the predicted probability of recurrence, which was interpreted as a continuous recurrence-risk score rather than a binary clinical classifier. Both Trained models, ALL and stacked ensemble, were then evaluated internally and on external data to assess generalization to an independent cohort.

### Repeated cross-validation and internal risk-score analysis

Internal model stability was assessed using repeated stratified 5-fold cross-validation with different random seeds at 100 iterations. Within each outer fold, feature selection, preprocessing, model tuning, and model fitting were performed using training samples only, and predicted probabilities were generated for held-out samples. Out-of-fold predicted probabilities were interpreted as recurrence-risk scores for RNA, copy-number, mutation, clinical, ALL, and stacked models. Patient-level recurrence-risk scores were then aggregated across iterations using the median predicted probability. Median risk scores were used for Kaplan–Meier analysis after dichotomization at the model-specific median and for Cox proportional hazards modeling as continuous predictors. Multivariable Cox models were adjusted for age and treatment covariates. Model stability was summarized by the number and proportion of iterations with significant log-rank or Cox associations.

### External validation

External validation was performed using publicly available METABRIC breast cancer molecular and clinical data obtained from GDC. METABRIC RNA expression, copy-number, mutation, immune/TME, and clinical data were processed to reproduce the feature classes used during model training. Validation features were determined from training feature-selection outputs and assembled into subtype-specific matrices matching the selected training features. Missing features after alignment were filled with 0. For each PAM50 subtype, the corresponding trained ALL and stacked ensemble recurrence-risk models were applied to the aligned METABRIC validation matrix. Model outputs were predicted probabilities of recurrence, which were interpreted as external recurrence-risk scores. External risk-score performance was evaluated using METABRIC recurrence-free survival outcome annotations. Predicted probabilities were analyzed as continuous risk scores and after stratification by prespecified percentile cutoffs, either median-based high- and low-risk groups or 75 and 25 percentile cutoffs. Kaplan–Meier analysis with log-rank testing was used to compare outcome distributions between risk groups, and Cox proportional hazards regression was used to estimate hazard ratios for model-derived risk scores.

### Data Availability

To comply with data access regulations, these sequencing data is available under controlled access; requests can be made to the corresponding author. All processed data inputs, scripts for model training and assessment, and, final models are available on GitHub at https://github.com/GavinHaLab/BRAVO_Recurrance_Risk_modeling for reproducibility.

## METHODS

### Overall study design

To discover and externally validate subtype-specific recurrence-risk signatures, we analyzed three subtype-defined discovery cohorts from the Breast Cancer Risk Factors and Various Outcomes (BRAVO) breast cancer cohort and one independent external validation cohort, METABRIC. The discovery cohorts consisted of primary breast tumors stratified by PAM50 subtype into Basal-like, Luminal A, and Luminal B groups. PAM50 subtypes were derived using the Genefu R package. Recurrent and non-recurrent tumors were represented at relatively similar proportions in the Basal-like cohort (71 recurrent and 72 non-recurrent tumors), Luminal A cohort (36 recurrent and 61 non-recurrent tumors), and Luminal B cohort (52 recurrent and 48 non-recurrent tumors).

We used the BRAVO cohort for discovery because it is highly annotated and includes manually curated medical record data, allowing detailed ascertainment of recurrence outcomes, recurrence-free survival time, primary treatments, demographics, clinical characteristics, and tumor features. Details of this cohort have been described previously^8,155,156^ Briefly, women 20-69 years of age with newly diagnosed first primary invasive stage I-III breast cancer were identified through the Cancer Surveillance System, the NCI-funded population-based Surveillance, Epidemiology, and End Results cancer registry serving western Washington state. All study procedures were reviewed and approved by the Fred Hutchinson Cancer Center Institutional Review Board and were conducted in accordance with U.S. Common Rule guidelines. Informed consent was obtained from all living participants, and eligible deceased participants were enrolled through an IRB-approved waiver of consent, reducing survival bias by allowing tumor tissue collection and medical record review for deceased patients.

Power calculations were performed separately for each subtype-specific discovery cohort based on the number of recurrent and non-recurrent tumors available for analysis. Using a one-sided test of AUC > 0.50 at α = 0.05, the Basal-like cohort had approximately 99.7% power to detect a discriminatory biomarker with AUC = 0.70 and 93.8% power to detect a biomarker with AUC = 0.65. The Luminal A cohort had approximately 96.0% and 80.0% power to detect biomarkers with AUC values of 0.70 and 0.65, respectively. The Luminal B cohort had approximately 97.8% and 84.1% power to detect biomarkers with AUC values of 0.70 and 0.65, respectively. For binary biomarkers with high specificity, AUC values of 0.70 and 0.65 correspond approximately to sensitivities of 40% and 30%, respectively.

External validation was performed in METABRIC, which was selected because it includes publicly available molecular data, PAM50 subtype annotations, clinical follow-up, and recurrence-associated survival outcomes. Final subtype-specific models were locked before application to METABRIC. No METABRIC samples were used for feature selection, model training, hyperparameter tuning, or model revision. Model-derived recurrence-risk scores were generated for METABRIC tumors within each corresponding subtype and evaluated for association with recurrence-free survival using Kaplan–Meier analysis and Cox proportional hazards regression.

### RNA and Targeted Panel library preparation and sequencing

Tumor total RNA was isolated from FFPE specimens using the AllPrep DNA/RNA/miRNA Kit (Qiagen). RNA sequencing was performed using an exome-capture transcriptome platform (Agilent SureSelect Exome V4) following standard protocols. DNA sequencing was performed using a targeted oncology panel, Roche Onco1500 v4; OncoPanelV4 or, Roche Onco1500 v2; OncoPanelV2. All sequencing was performed on FFPE-derived nucleic acids. All samples were sequenced on the Illumina HiSeq 2000 or HiSeq 2500 in paired-end mode.

### RNA-sequencing preprocessing, quality control, and differential expression analysis

Raw paired-end RNA-sequencing reads were assessed with FastQC, adapter- and quality-trimmed using Trim Galore, and filtered for ribosomal RNA using BBDuk. Filtered reads were quasi-mapped and quantified with Salmon against a GENCODE release 41 transcriptome index. Salmon files were imported using tximport, with transcript-to-gene mappings derived from the GENCODE v41 GTF and converted to gene-level count and abundance matrices. The TPM matrix was retained for downstream expression-based analyses and feature extraction.

RNA-seq quality control was performed using expression-based metrics, including total transcript counts, library complexity, and number of detected genes. Samples failing two or more QC metrics were excluded, resulting in removal of 23 RNA-seq samples from downstream analyses. Any remaining ribosomal genes were identified using an HGNC ribosomal gene list and removed from the count matrix prior to differential expression analysis as an additional measure to remove rRNA. RNA-seq quality control metrics, including sequencing (from Multiqc and BBDuk) and expression-based metrics (mentioned above), are provided in **Supplemental Table 9 and Supplemental Table 10**.

Differential expressions was performed separately within each PAM50 subtype using limma-voom. For each subtype, raw gene-level counts were normalized using edgeR TMM normalization, transformed using voom, and modeled with limma linear models followed by empirical Bayes moderation. The design matrix included recurrence status only, and the contrast was defined as recurrent versus non-recurrent disease. Genes were retained for testing if they had a total count of ≥50 and had ≥10 counts in at least 10% of samples. Differentially expressed genes were defined as those with nominal P < 0.05, absolute log2 fold change > 0.5, and base mean expression > 50. Gene-level Cox proportional hazards models were also fit using recurrence as the event and adjusting for age and treatment; genes were considered Cox-associated if P < 0.05. Genes were retained as candidate RNA model features if they met either the limma differential-expression criteria or the adjusted Cox regression criteria.

### Gene set enrichment analysis

Preranked GSEA was performed separately within each PAM50 subtype using recurrence-associated differential expression results. Genes were ranked by multiplying log2 fold change by the negative log10 nominal P value, generating a signed statistic in which positive values corresponded to genes increased in recurrent tumors and negative values corresponded to genes increased in non-recurrent tumors. Ranked gene lists were tested against KEGG and Hallmark gene-set collections. Pathways were considered enriched if nominal P < 0.05 and FDR q < 0.25.

For heatmap visualization, pathways were prioritized by significance, normalized enrichment score, and concordance across subtypes, then separated into intrinsic tumor-associated and immune-associated gene-set groups. Intrinsic and immune pathways were hierarchically clustered separately using subtype-specific normalized enrichment scores. Cross-hatched pathways did not meet the prespecified enrichment threshold but were retained to show directional concordance across subtypes; full NES, nominal P values, and FDR q values are provided in Supplementary Table 2.

### Gene set variation analysis

Sample-level pathway activity was quantified using GSVA on log2(TPM + 1) expression values for Hallmark, KEGG, and Reactome gene sets. Analyses were performed separately within each PAM50 subtype. GSVA scores were compared between recurrent and non-recurrent tumors, and pathway-level Cox proportional hazards models were fit using recurrence as the event, adjusting for age and treatment. Pathways were considered recurrence-associated if nominal P < 0.05 and FDR q < 0.25 in the group comparison or adjusted Cox analysis. Hazard ratios were used to indicate the direction and magnitude of recurrence-associated pathway activity. GSVA scores were incorporated as RNA pathway-level features in downstream recurrence-risk models.

### Molecular signature analysis

Published molecular signatures were scored from TPM-derived RNA expression values using either geometric mean log2TPM or median log2(TPM + 1) values across signature genes. Signatures included innate and adaptive enriched tumors, CLINSAR, and several Epithelial to Mesenchymal Transition / Plasticity signatures.^27–31,157–163^ Analyses were performed separately within each PAM50 subtype. Signature scores were compared between recurrent and non-recurrent tumors with Mann–Whitney U and were also evaluated using Cox proportional hazards models with recurrence as the event, adjusting for age and treatment. Signature scores were also binarized (median) and compared between recurrent and non-recurrent tumors with Fisher exact and Cox proportional hazards models with recurrence as the event, adjusting for age and treatment. Signatures were retained as candidate model features if either the group comparison or adjusted Cox regression met nominal P < 0.05.

### Tumor microenvironment analysis

Tumor microenvironment features were evaluated using RNA expression (log2(TPM + 1)) with CIBERSORT-derived immune cell fractions, EcoTyper cell-state abundances, and TIDE immune dysfunction/exclusion scores.^62,106,164^ CIBERSORT and EcoTyper Carcinoma Types were analyzed across PAM50 subtypes to assess overall subtype-associated immune composition differences. CIBERSORT, EcoTyper Carcinoma Types and EcoTyper Cell States, and TIDE scores were separately assessed within each subtype to evaluate the recurrence-associated tumor microenvironment. Within-subtype comparisons between recurrent and non-recurrent tumors were performed using the Mann–Whitney U test. Cox proportional hazards models were also fit using recurrence as the event, adjusting for age and treatment. Features were considered recurrence-associated if nominal P < 0.05 by either Mann–Whitney U testing or adjusted Cox regression. For heatmap visualization, CIBERSORT features were z-scored (within subtype) and differences in mean subtype-scaled feature abundance between recurrent and non-recurrent tumors were calculated. Hierarchical clustering was used to organize CIBERSORT features in heatmaps.

### Targeted panel alignment and preprocessing

Paired-end targeted-panel FASTQ files were processed using a Snakemake workflow. Reads were aligned to the reference genome using BWA-MEM, then converted to BAM format using SAMtools. BAM files were coordinate-sorted with SAMtools, and duplicate reads were marked using Picard MarkDuplicates. Base-quality score recalibration was performed using GATK BaseRecalibrator with known polymorphic sites specified in the workflow configuration, followed by ApplyBQSR to generate final recalibrated BAM files and indexes. Alignment quality metrics, duplicate metrics, and insert-size metrics were collected using Picard. Genomic quality-control metrics for targeted-panel sequencing including Picard HS, insert size and alignment metrics are in **Supplementary Table 11**.

### Targeted panel mutation calling and annotation

Somatic variants were called from targeted-panel tumor–normal BAM files using GATK Mutect2 and Strelka2 with matched-normal filtering. Variants were retained for downstream analysis only if they were identified by both callers. Consensus variants were annotated with ANNOVAR against hg38 using gene, population-frequency, cancer, clinical, and functional annotation databases, including RefGene, COSMIC, ClinVar, dbNSFP, InterVar, gnomAD, dbSNP, and 1000 Genomes resources. ANNOVAR annotation was performed with VCF input and nearest-gene annotation using a 5-kb window.

Consensus somatic variants were filtered to reduce germline contamination, low-confidence calls, and FFPE-associated artifacts. Variants were required to have AF_popmax <0.10, tumor and normal depth ≥10 reads, tumor variant allele fraction ≥0.02, normal variant allele fraction ≤0.10, tumor alternate depth ≥3 reads, and normal reference depth ≥8 reads. SOBDetector was applied to assess FFPE-associated strand-orientation artifacts, and variants predicted to represent FFPE artifacts were excluded before downstream mutation analyses.^165^ Because FFPE-associated artifacts are enriched for low-frequency C>T/G>A transitions, C>T and G>A variants were subjected to an additional VAF threshold. Consistent with prior FFPE filtering strategies, C>T/G>A variants were retained only if tumor VAF was ≥0.15 and they passed SOBDetector artifact filtering.^165,166^

### Pathway tumor mutational burden analysis

Pathway-level mutation features were generated from filtered consensus somatic SNVs. For each sample, pathway tumor mutational burden was calculated using an approach adapted from PathwayTMB, in which gene-level mutation density was calculated for each pathway gene and then summarized across the pathway:^167^

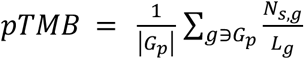

Where *s* is the sample, *p* is the pathway, *G_p_* is the set of genes in pathway *p*, *n_s,g_*, is the number of filtered SNVs in gene *g* for sample *s*, and *L_g_* is the coding length of gene *g*.

In parallel, binary pathway mutation features were generated to indicate whether any gene in a pathway was mutated in a given sample.^70^ Both continuous pathway TMB values and binary pathway mutation indicators were evaluated separately within each PAM50 subtype. Recurrence-associated differences were assessed using distributional testing between recurrent and non-recurrent tumors and Cox proportional hazard regression with recurrence as the event, adjusting for age and treatment. For overall significance analysis, pathways were pruned for redundancy by determining the overall overlap in genes between pathways and dropping pathways when overlap was greater than 0.75 of genes. Pathway mutation features were retained for downstream modeling if either the distributional comparison or adjusted Cox regression met nominal P < 0.05.

### Copy-number analysis

Copy-number profiles were generated from targeted-panel MiOncoSeq data using a modified CNVkit–ichorCNA pipeline. Because all samples were from female patients, chromosome X was retained and chromosome Y was excluded from downstream copy-number analyses. Gene-level copy-number values were normalized by sample ploidy and converted to copy-number deviation by subtracting 1, such that 0 represented copy-number neutral, positive values represented gains, and negative values represented losses. Gene-level gains and losses were defined using copy-number deviation thresholds of ≥0.3 and ≤−0.3, respectively. Cytoband-level copy-number features were generated by mapping genes to genomic coordinates and assigning genes to cytobands. For each sample and cytoband, binary amplification/deletion calls were calculated. Cytoband amplification or deletion was called when at least 60% of mapped genes in the band met the corresponding gain or loss threshold, and cytobands with fewer than three mapped genes were excluded. Arm-level and genome-level copy-number burden features were also calculated, including percent arm amplified, percent arm deleted, and percent genome altered. Arm-level events were binarized as present when the corresponding percent alteration was greater than 80%. To evaluate pathway-level copy-number patterns, GSVA was performed on the gene-level copy-number deviation matrix using variance-filtered pathway gene sets. This generated sample-level copy-number pathway activity scores, which were analyzed within each PAM50 subtype. Each copy number feature type was compared between recurrent and non-recurrent tumors within each subtype and were also evaluated using Cox proportional hazards regression with recurrence as the event, adjusting for age and treatment. Copy-number features were retained for downstream modeling if they met nominal P < 0.05 by either recurrence-group testing or adjusted Cox regression. Gene-level, cytoband-level, arm-level, genome-level, and CN-GSVA pathway-level copy-number features were included as candidate inputs for downstream recurrence-risk modeling.

### Risk Model Feature Selection

Feature selection was performed independently within each PAM50 subtype using the following sequential filtering: variance, correlation with recurrence, recurrence-association testing, and redundancy-pruning **(Fig.4A)**. Candidate RNA, copy-number, mutation, and molecular-signature features were first filtered using label-free variance criteria to remove low-variability predictors keeping only the top 40% of features. Features passing this step were then screened using a supervised feature–label correlation filter (r > 0.15), with recurrence encoded as a binary outcome. Remaining features were tested for recurrence association using two-sided Mann–Whitney U tests for continuous features and Fisher’s exact tests for binary features. In parallel, feature-wise Cox proportional hazards models were fit using recurrence as the event and adjusting for age and treatment covariates. Features were retained if they met nominal P < 0.05 by at least one recurrence-association test: Mann–Whitney U, Fisher’s exact test, or adjusted Cox regression. Selected features were then pruned for redundancy within molecular feature blocks using feature–feature correlation. When groups of highly correlated features exceeded the prespecified correlation threshold (0.90), a single representative feature was retained based on the strongest recurrence-associated evidence, prioritizing adjusted Cox regression results when available. This produced final non-redundant, subtype-specific feature matrices for downstream recurrence-risk modeling.

### Model training and model selection

Subtype-specific recurrence-risk classifiers were trained using the final post-feature-selection features. For each PAM50 subtype, recurrence status was encoded as a binary outcome, with recurrent tumors assigned to class 1 and non-recurrent tumors assigned to class 0. Candidate predictors included four feature blocks: RNA-derived features (including immune/TME features, molecular signature scores), copy-number features, mutation-derived features, and clinical covariates. Model-family selection was performed independently within each subtype and feature block using 5-fold cross-validation with ROC AUC as the optimization metric. Candidate model families were evaluated separately for RNA, copy-number, mutation, clinical, and concatenated ALL feature matrices. For each subtype/block combination, the model family with the highest mean cross-validated ROC AUC was selected and fixed for final training. The final locked model families for each stacked block (RNA, CNV, MUT & CLINICAL) as determined by 5-fold cv were Gradient Boosting for Basal RNA, mutation, and ALL features, ExtraTrees for Basal copy number, and L2-penalized logistic regression for Basal clinical features; LinearSVC for Luminal A RNA and clinical features, Random Forest for Luminal A copy-number and SVM for mutation features, and L2-penalized logistic regression for Luminal A ALL features; and Gradient Booting for Luminal B RNA features, Random Forest for Luminal B copy-number features, ElasticNet penalized logistric regression for Luminal B mutation features, and L1-penalized logistic regression for Luminal B clinical features and ALL features. Final hyperparameter tuning was then performed using GridSearchCV restricted to the selected model family, and the best model was refit using all available training samples within that subtype. Continuous molecular features were standardized using z-score scaling within the training data. Binary pathways mutations, clinical variables, discrete copy-number calls, and other binary/discrete features were passed through without scaling. Class imbalances were addressed during training using recurrent-class oversampling and class-weight tuning when supported by the model family. The two main integrated modeling strategies trained were the ALL model, trained as a single classifier using the full concatenated feature matrix across all retained molecular feature blocks, and a stacked ensemble model, trained using the feature block-level classifiers (as listed above). For the stacked model, each block-level classifier generated predicted probabilities of recurrence, and these probabilities were used as meta-features for either an L1 or L2-regularized logistic regression meta-model (Basal: L1, Luminal A: L1, Luminal B: L2). Final hyperparameter tuning of the meta model was performed using GridSearchCV. The primary model output (both ALL and stacked ensemble) was the predicted probability of recurrence, which was interpreted as a continuous recurrence-risk score rather than a binary clinical classifier. Both Trained models, ALL and stacked ensemble, were then evaluated internally and on external data to assess generalization to an independent cohort.

### Repeated cross-validation and internal risk-score analysis

Internal model stability was assessed using repeated stratified 5-fold cross-validation with different random seeds at 100 iterations. Within each outer fold, feature selection, preprocessing, model tuning, and model fitting were performed using training samples only, and predicted probabilities were generated for held-out samples. Out-of-fold predicted probabilities were interpreted as recurrence-risk scores for RNA, copy-number, mutation, clinical, ALL, and stacked models. Patient-level recurrence-risk scores were then aggregated across iterations using the median predicted probability. Median risk scores were used for Kaplan–Meier analysis after dichotomization at the model-specific median and for Cox proportional hazards modeling as continuous predictors. Multivariable Cox models were adjusted for age and treatment covariates. Model stability was summarized by the number and proportion of iterations with significant log-rank or Cox associations.

### External validation

External validation was performed using publicly available METABRIC breast cancer molecular and clinical data obtained from GDC. METABRIC RNA expression, copy-number, mutation, immune/TME, and clinical data were processed to reproduce the feature classes used during model training. Validation features were determined from training feature-selection outputs and assembled into subtype-specific matrices matching the selected training features. Missing features after alignment were filled with 0. For each PAM50 subtype, the corresponding trained ALL and stacked ensemble recurrence-risk models were applied to the aligned METABRIC validation matrix. Model outputs were predicted probabilities of recurrence, which were interpreted as external recurrence-risk scores. External risk-score performance was evaluated using METABRIC recurrence-free survival outcome annotations. Predicted probabilities were analyzed as continuous risk scores and after stratification by prespecified percentile cutoffs, either median-based high- and low-risk groups or 75 and 25 percentile cutoffs. Kaplan–Meier analysis with log-rank testing was used to compare outcome distributions between risk groups, and Cox proportional hazards regression was used to estimate hazard ratios for model-derived risk scores.

### Data Availability

Sequencing data generated in this study is available upon formal request. To comply with data access regulations, these data are available under controlled access; requests can be made to the corresponding author. All inputs and scripts for model training and assessment are available on GitHub at https://github.com/GavinHaLab/BRAVO_Recurrance_Risk_modeling for reproducibility.

## AUTHOR CONTRIBUTIONS

Conceptualization: C.L. G.H. E.C.

Methodology: E.C.

Software: E.C.

Formal Analysis: E.C. P.C.

Investigation: E.C.

Resources: G.H. C.L.

Data Curation: C.L.

Writing – Original Draft: E.C.

Writing – Review & Editing: E.C. G.H. C.L.

Visualization: E.C.

Supervision: G.H. C.L.

Funding Acquisition: G.H. C.L.

## ACKNOWLEDGEMENTS

We thank the MI-ONCOSEQ team at the University of Michigan for contributing to data generation, processing and analysis support of targeted panel and RNA sequencing data for this study. Specifically, we acknowledge Yi-Mi Wu, Dan Robinson, Xuhong Cao, Marcin Cieslik, and Arul Chinnaiyan for their contributions and support.

## SUPPLEMENTAL FIGURES

**Supplementary Figure 1.**
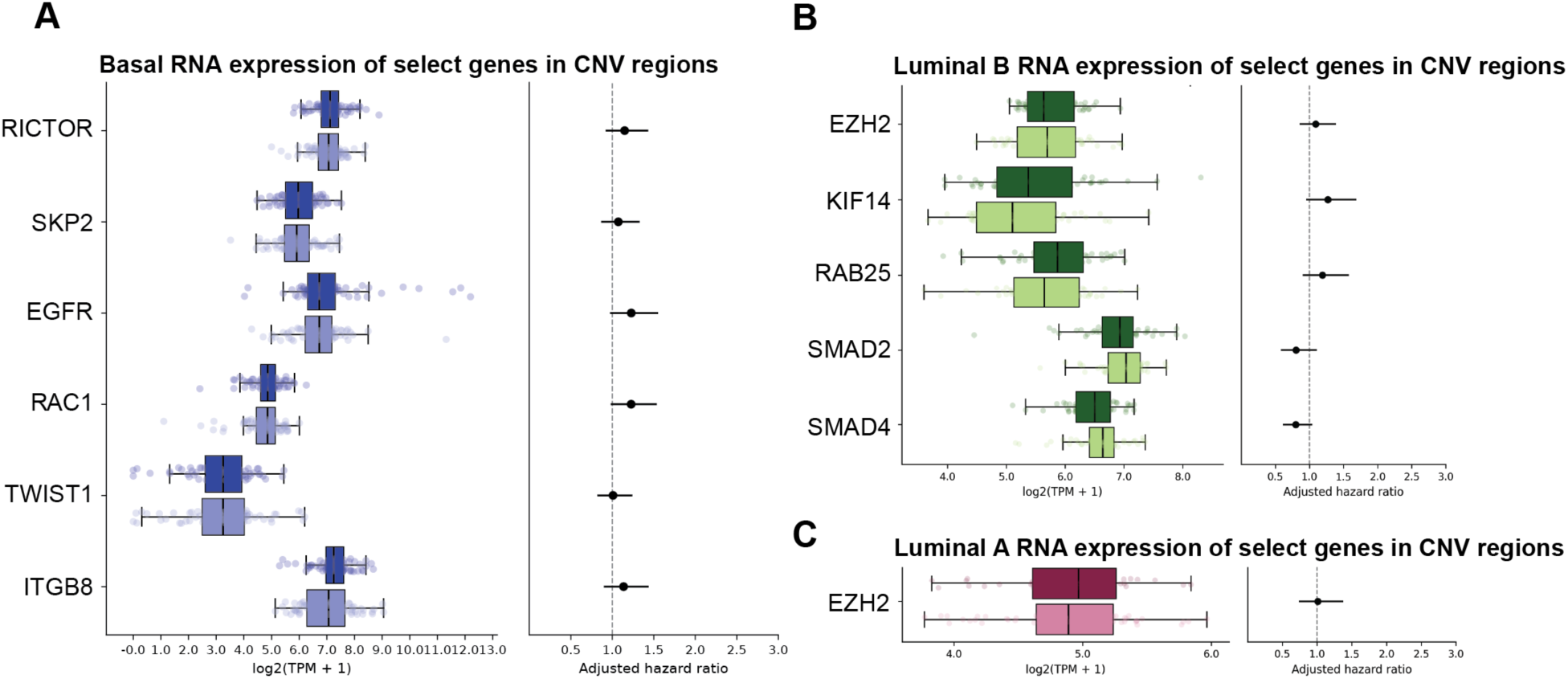
Gene expression of recurrence-associated copy-number region genes across breast cancer subtypes. (a) Basal-like recurrence-associated gene expression. Box plots show log2(TPM) expression distributions of selected genes from recurrence-associated copy-number regions (RICTOR, SKP2, EGFR, RAC1, TWIST1, ITGB8) in recurrent and non-recurrent tumors (left), with corresponding adjusted hazard ratios for recurrence-free survival by Cox regression (right). Asterisks indicate statistical significance by Mann–Whitney U test. (b) Luminal B recurrence-associated gene expression, displayed as in (a), for selected genes from recurrence-associated copy-number regions (KIF14, RAB25, EZH2). (c) Luminal A recurrence-associated gene expression, displayed as in (a), for selected genes from recurrence-associated copy-number regions (EZH2).

**Supplementary Figure 2.**
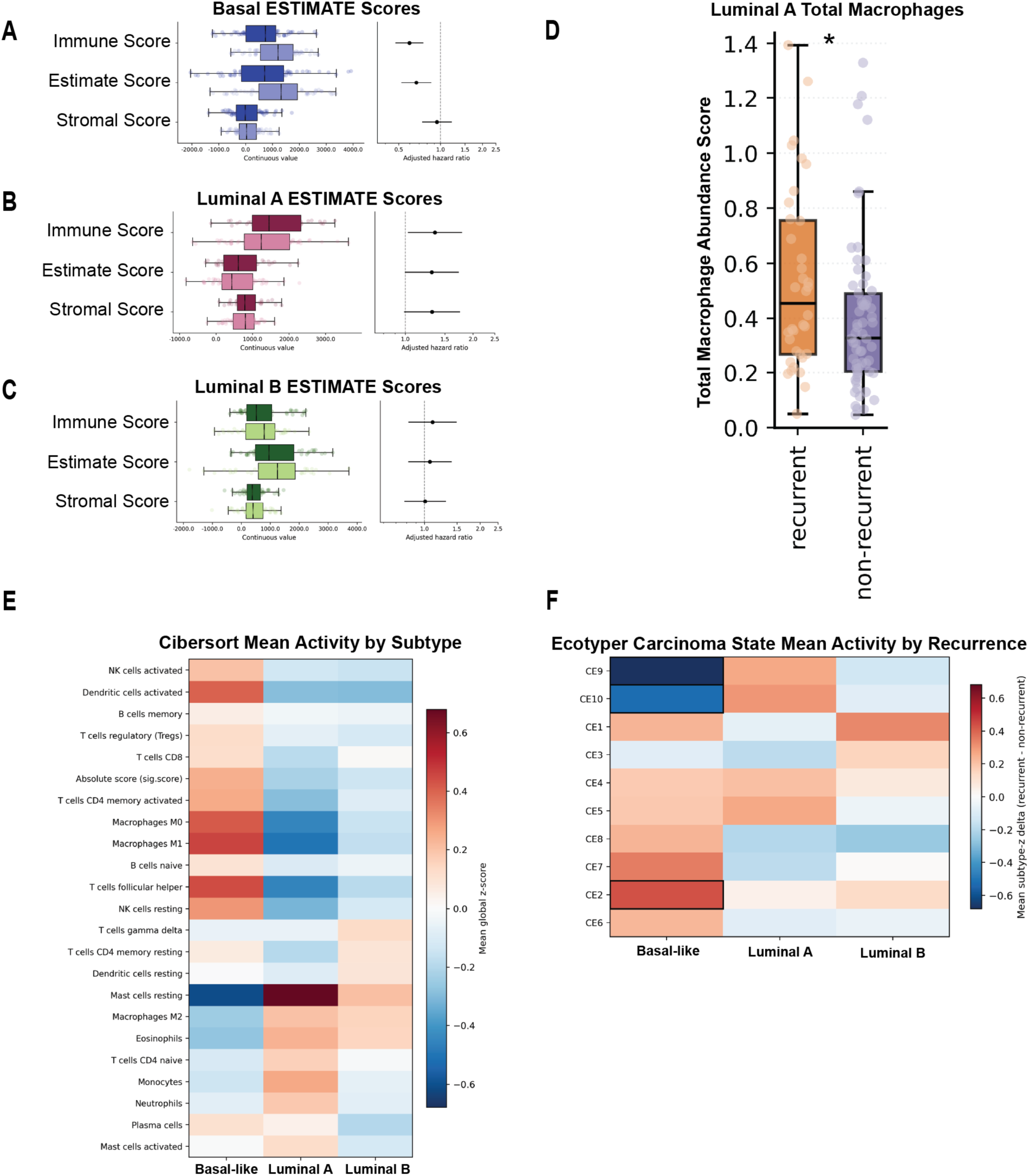
Tumor microenvironment characterization across breast cancer subtypes. (a–c) ESTIMATE scores for Basal-like (a), Luminal A (b), and Luminal B (c) subtypes. Box plots show immune score, stromal score, and ESTIMATE score distributions in recurrent and non-recurrent tumors (left), with corresponding adjusted hazard ratios for recurrence-free survival by Cox regression (right). CIBERSORT abundance score of total macrophages (M0, M1, M2) for Luminal A recurrent and non-recurrent tumors. Asterisks indicate statistical significance by MWU. p < 0.05. (e) Heatmap of mean CIBERSORT absolute immune cell type scores across Basal-like, Luminal A, and Luminal B subtypes. Color reflects mean cell type abundance per subtype, with warm colors indicating higher abundance and cool colors indicating lower abundance. (f) Heatmap of mean EcoTyper carcinoma cell state activity by recurrence status across Basal-like, Luminal A, and Luminal B subtypes. Color reflects the mean subtype-z delta score (recurrent minus non-recurrent), with warm colors indicating higher activity in recurrent tumors and cool colors indicating higher activity in non-recurrent tumors. Outlined states indicate statistically significant differences.

**Supplementary Figure 3.**
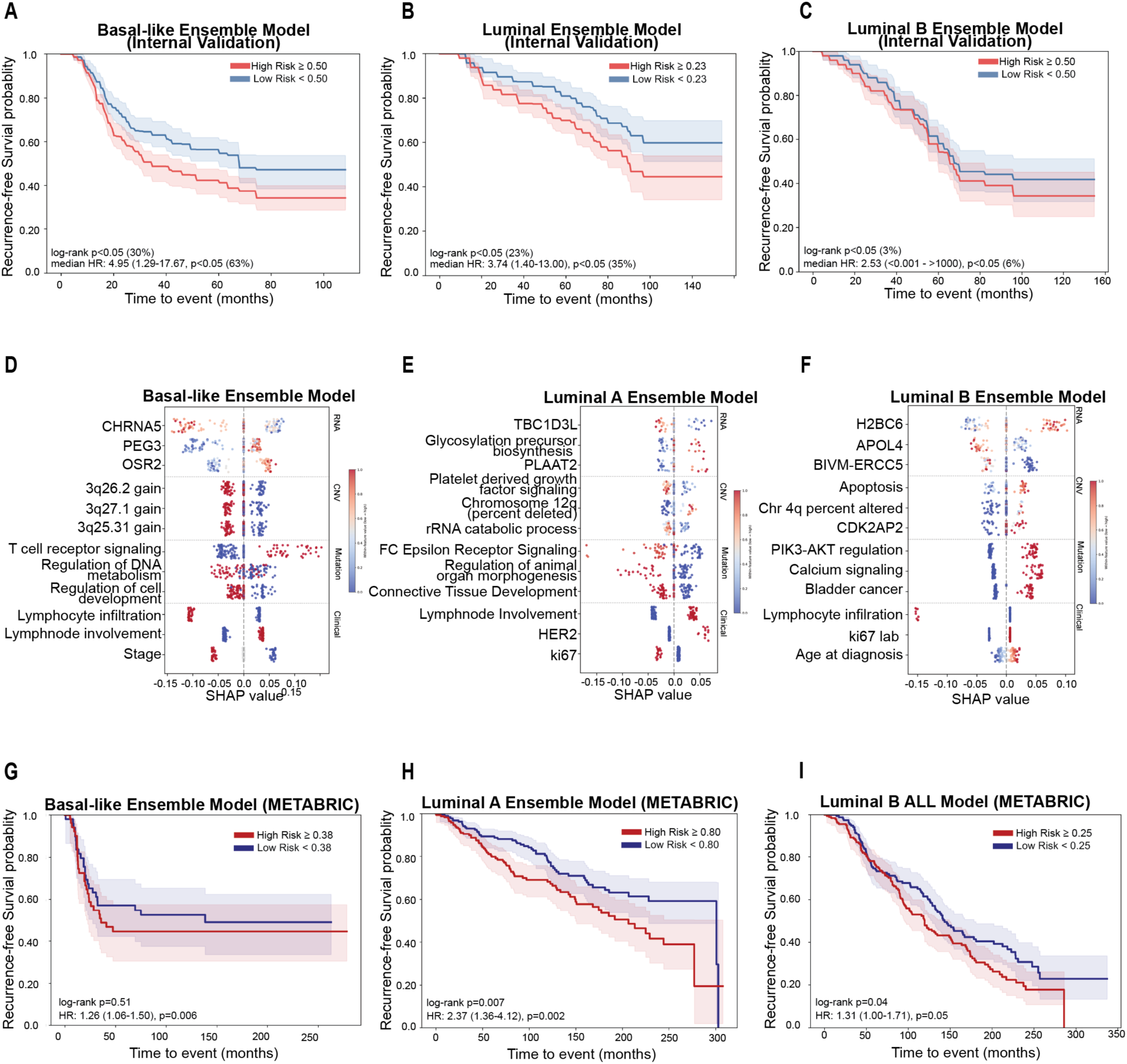
Internal and external validation of alternative subtype-specific recurrence-risk models. (a–c) Internal validation Kaplan-Meier curves for recurrence-free survival for the Basal-like Ensemble model (a), Luminal A Ensemble model (b), and Luminal B Ensemble model (c). Percent of log-rank p-values <0.05, median hazard ratios with 95% confidence intervals, and median high risk / low risk cut offs are shown. Shaded regions represent 95% confidence intervals. (d–f) SHAP feature importance plots for the Basal-like Ensemble model (d), Luminal A Ensemble model (e), and Luminal B ALL model (f). Points represent individual samples separated by data modality (RNA, CNV, mutation, clinical), with SHAP values indicating direction and magnitude of contribution to the risk score. (g–i) External validation Kaplan-Meier curves in the METABRIC cohort for the Basal-like Ensemble model (g), Luminal A Ensemble model (h), and Luminal B ALL model (i). Log-rank p-values and hazard ratios based on continuous (Luminal A & Basal) or binarized (Luminal B) recurrence risk scores with 95% confidence intervals are shown.

## SUPPLEMENTAL TABLE LEGENDS

**Supplementary Table 1**. **Clinical and metadata summary by subtype and recurrence status.** Basal-like, Luminal A, and Luminal B tumors are summarized overall and by recurrence status. Categorical variables are shown as counts and percentages, and continuous variables are shown as mean and median values. P values compare recurrent and non-recurrent tumors within each subtype using the statistical test listed in the table.

**Supplementary Table 2. Subtype-specific RNA-seq differential expression and pathway analysis results.** This supplemental data contains full RNA-seq results comparing recurrent and non-recurrent tumors within Basal-like, Luminal A, and Luminal B subtypes. Sheets include gene-level limma differential expression results, positively and negatively enriched GSEA pathways, and recurrence-associated custom gene-set/signature analyses with group-comparison, survival, Cox model, and FDR-adjusted statistics.

**Supplementary Table 3. Copy-number recurrence-association results at cytoband and chromosome-arm levels.** This supplemental data contains subtype-stratified copy-number analyses comparing recurrent and non-recurrent Basal-like, Luminal A, and Luminal B tumors. For each subtype, cytoband-level results summarize amplification and deletion event frequencies, and Cox proportional hazards estimates. Chromosome-arm–level sheets summarize arm-level amplifications and deletions, with Cox model associations.

**Supplementary Table 4. Subtype-specific pathway-level tumor mutational burden results.** This supplemental data contains pathway-level tumor mutational burden analyses comparing recurrent and non-recurrent tumors within Basal-like, Luminal A, and Luminal B subtypes. Sheets include full pTMB results, redundancy-mapping outputs, and final pruned non-redundant pathway results.

**Supplementary Table 5. Subtype-specific immune composition scores.** This supplemental data contains immune composition analyses for Basal-like, Luminal A, and Luminal B tumors, including ESTIMATE stromal, immune and purity scores, CIBERSORT immune-cell fractions, recurrence comparisons of immune-cell compositions, subtype-level statistical tests, and heatmap summary values.

**Supplementary Table 6. Subtype-specific EcoTyper cell-state and carcinoma-state results.** This supplemental data contains EcoTyper-derived cell-state and carcinoma-state abundance analyses for Basal-like, Luminal A, and Luminal B tumors, including subtype-specific recurrence comparisons, carcinoma-state summaries across subtypes, and statistical tests of carcinoma-state enrichment.

**Supplementary Table 7. Subtype-specific TIDE immune-evasion and T-cell–inflamed signature results.** This spreadsheet contains subtype-level TIDE-derived immune-evasion analysis and T-cell–inflamed signature analysis for Basal-like, Luminal A, and Luminal B tumors.

**Supplementary Table 8. Subtype-specific recurrence-risk model feature selection, model selection, internal validation results and external validation results.** This supplemental data contains model-selected features, model-family selection from cross validation ALL model and ensemble blocks, and internal cross-validation results for Basal-like, Luminal A, and Luminal B recurrence-risk models, including Cox and log-rank performance summaries across iterations and Cox hazard ratios for external validation performance.

**Supplementary Table 9 and 10. RNA-seq quality-control metrics by sample.** These supplemental data tables contain RNA-seq quality-control metrics for profiled samples, including sequencing- and expression-based measures.

**Supplementary Table 11. Targeted-panel sequencing quality-control metrics by sample.** This supplemental data contains targeted-panel sequencing metrics for profiled samples, including hybrid-selection metrics, insert-size distributions, and alignment metrics used to evaluate sequencing quality and sample inclusion.

## REFERENCES

1. Breast Cancer Statistics | How Common Is Breast Cancer? https://www.cancer.org/cancer/types/breast-cancer/about/how-common-is-breast-cancer.html.

2. Liu, M. C. et al. PAM50 gene signatures and breast cancer prognosis with adjuvant anthracycline- and taxane-based chemotherapy: correlative analysis of C9741 (Alliance). Npj Breast Cancer 2, 15023 (2016).

3. Ribelles, N. et al. Pattern of recurrence of early breast cancer is different according to intrinsic subtype and proliferation index. Breast Cancer Res. BCR 15, R98 (2013).

4. Visser, K. E. de & Joyce, J. A. The evolving tumor microenvironment: From cancer initiation to metastatic outgrowth. Cancer Cell 41, 374–403 (2023).

5. Mullins, R., Pal, A., Barrett, T. F., Neal, M. E. H. & Puram, S. V. Epithelial-mesenchymal plasticity in tumor immune evasion. Cancer Res. 82, 2329–2343 (2022).

6. Tekpli, X. et al. An independent poor-prognosis subtype of breast cancer defined by a distinct tumor immune microenvironment. Nat. Commun. 10, 5499 (2019).

7. Sharma, A. et al. Comprehensive multi-omics analysis of breast cancer reveals distinct long-term prognostic subtypes. Oncogenesis 13, 22 (2024).

8. Li, C. I. et al. Cancer Cell Intrinsic and Immunologic Phenotypes Determine Clinical Outcomes in Basal-like Breast Cancer. Clin. Cancer Res. 27, 3079–3093 (2021).

9. Li, J., Wu, J. & Han, J. Analysis of Tumor Microenvironment Heterogeneity among Breast Cancer Subtypes to Identify Subtype-Specific Signatures. Genes 14, 44 (2022).

10. Vieira, A. F. & Schmitt, F. An Update on Breast Cancer Multigene Prognostic Tests—Emergent Clinical Biomarkers. Front. Med. 5, (2018).

11. Kittaneh, M., Montero, A. J. & Glück, S. Molecular Profiling for Breast Cancer: A Comprehensive Review. Biomark. Cancer 5, BIC.S9455 (2013).

12. Hsieh, T.-Y., Huang, C.-C. & Tseng, L.-M. Multi-gene expression assays in breast cancer: a literature review. Transl. Cancer Res. 14, 6092–6101 (2025).

13. Brogna, M. R. et al. Evaluation and Comparison of Prognostic Multigene Tests in Early-Stage Breast Cancer: Which Is the Most Effective? A Literature Review Exploring Clinical Utility to Enhance Therapeutic Management in Luminal Patients. Mol. Carcinog. 64, 789–800 (2025).

14. Sparano, J. A. et al. Prospective Validation of a 21-Gene Expression Assay in Breast Cancer. N. Engl. J. Med. 373, 2005–2014 (2015).

15. Bartlett, J. M. S. et al. Comparative survival analysis of multiparametric tests—when molecular tests disagree—A TEAM Pathology study. NPJ Breast Cancer 7, 90 (2021).

16. Fan, C. et al. Building prognostic models for breast cancer patients using clinical variables and hundreds of gene expression signatures. BMC Med. Genomics 4, 3 (2011).

17. Wang, D., Gao, S., Qian, H., Yuan, P. & Zhang, B. Prognostic Value of Copy Number Alteration Burden in Early-Stage Breast Cancer and the Construction of an 11-Gene Copy Number Alteration Model. Cancers 14, 4145 (2022).

18. Jin, X., Yan, J., Chen, C., Chen, Y. & Huang, W.-K. Integrated Analysis of Copy Number Variation, Microsatellite Instability, and Tumor Mutation Burden Identifies an 11-Gene Signature Predicting Survival in Breast Cancer. Front. Cell Dev. Biol. 9, 721505 (2021).

19. Curtis, C. et al. The genomic and transcriptomic architecture of 2,000 breast tumours reveals novel subgroups. Nature 486, 346–352 (2012).

20. Nolan, E., Lindeman, G. J. & Visvader, J. E. Deciphering breast cancer: from biology to the clinic. Cell 186, 1708–1728 (2023).

21. Sammut, S.-J. et al. Multi-omic machine learning predictor of breast cancer therapy response. Nature 601, 623–629 (2022).

22. Llinas-Bertran, A., Butjosa-Espín, M., Barberi, V. & Seoane, J. A. Multimodal data integration in early-stage breast cancer. The Breast 80, 103892 (2025).

23. Jiang, Z., Zhang, H., Gao, Y. & Sun, Y. Multi-omics strategies for biomarker discovery and application in personalized oncology. Mol. Biomed. 6, 115 (2025).

24. Bendani, H., Boumajdi, N., Belyamani, L. & Ibrahimi, A. Toward precision oncology: An integrative multi-omics approach for prognosis prediction and inferred immunotherapy responsiveness in breast cancer. Clin. Transl. Discov. 6, e70116 (2026).

25. Parker, J. S. et al. Supervised Risk Predictor of Breast Cancer Based on Intrinsic Subtypes. J. Clin. Oncol. 27, 1160–1167 (2009).

26. Pereira, B. et al. The somatic mutation profiles of 2,433 breast cancers refine their genomic and transcriptomic landscapes. Nat. Commun. 7, 11479 (2016).

27. Mak, M. P. et al. A Patient-Derived, Pan-Cancer EMT Signature Identifies Global Molecular Alterations and Immune Target Enrichment Following Epithelial-to-Mesenchymal Transition. Clin. Cancer Res. 22, 609–620 (2016).

28. Cheng, Q. et al. A signature of epithelial-mesenchymal plasticity and stromal activation in primary tumor modulates late recurrence in breast cancer independent of disease subtype. Breast Cancer Res. BCR 16, 407 (2014).

29. Gordian, E. et al. Transforming growth factor Î^2^induced epithelialtomesenchymal signature predicts metastasisfree survival in nonsmall cell lung cancer. Oncotarget 10, 810–824 (2019).

30. Chibon, F. et al. Validated prediction of clinical outcome in sarcomas and multiple types of cancer on the basis of a gene expression signature related to genome complexity. Nat. Med. 16, 781–787 (2010).

31. Xue, W. et al. Wnt/β-catenin-driven EMT regulation in human cancers. Cell. Mol. Life Sci. CMLS 81, 79 (2024).

32. Serrano, I., McDonald, P. C., Lock, F. E. & Dedhar, S. Role of the integrin-linked kinase (ILK)/Rictor complex in TGFβ-1-induced epithelial–mesenchymal transition (EMT). Oncogene 32, 50–60 (2013).

33. Ansari, M. N. et al. The mTORC2 subunit RICTOR drives breast cancer progression by promoting ganglioside biosynthesis through transcriptional and epigenetic mechanisms. PLOS Biol. 23, e3003362 (2025).

34. Yang, Q. et al. Acquisition of epithelial–mesenchymal transition is associated with Skp2 expression in paclitaxel-resistant breast cancer cells. Br. J. Cancer 110, 1958–1967 (2014).

35. Park, H. S. et al. High EGFR gene copy number predicts poor outcome in triple-negative breast cancer. Mod. Pathol. 27, 1212–1222 (2014).

36. ERK2 regulates epithelial-to-mesenchymal plasticity through DOCK10-dependent Rac1/FoxO1 activation | PNAS. https://www.pnas.org/doi/10.1073/pnas.1811923116.

37. Baugher, P. J., Krishnamoorthy, L., Price, J. E. & Dharmawardhane, S. F. Rac1 and Rac3 isoform activation is involved in the invasive and metastatic phenotype of human breast cancer cells. Breast Cancer Res. BCR 7, R965–974 (2005).

38. Kim, M. S. et al. MEST induces Twist-1-mediated EMT through STAT3 activation in breast cancers. Cell Death Differ. 26, 2594–2606 (2019).

39. Takasaka, N. et al. Integrin αvβ8–expressing tumor cells evade host immunity by regulating TGF-β activation in immune cells. JCI Insight 3, e122591.

40. Mitra, S. et al. Rab25 acts as an oncogene in luminal B breast cancer and is causally associated with Snail driven EMT. Oncotarget 7, 40252–40265 (2016).

41. Pariyar, M., Johns, A., Thorne, R. F., Scott, R. J. & Avery-Kiejda, K. A. Copy number variation in triple negative breast cancer samples associated with lymph node metastasis. Neoplasia 23, 743–753 (2021).

42. Jang, S.-H. et al. High EZH2 Protein Expression Is Associated with Poor Overall Survival in Patients with Luminal A Breast Cancer. J. Breast Cancer 19, 53–60 (2016).

43. Oshi, M. et al. The E2F Pathway Score as a Predictive Biomarker of Response to Neoadjuvant Therapy in ER+/HER2− Breast Cancer. Cells 9, 1643 (2020).

44. Liu, N. et al. SMAD4 is a potential prognostic marker in human breast carcinomas. Tumour Biol. J. Int. Soc. Oncodevelopmental Biol. Med. 35, 641–650 (2014).

45. Li, H. et al. Targeting of mTORC2 prevents cell migration and promotes apoptosis in breast cancer. Breast Cancer Res. Treat. 134, 1057–1066 (2012).

46. du Rusquec, P., Blonz, C., Frenel, J. S. & Campone, M. Targeting the PI3K/Akt/mTOR pathway in estrogen-receptor positive HER2 negative advanced breast cancer. Ther. Adv. Med. Oncol. 12, 1758835920940939 (2020).

47. Morrison Joly, M., et al. Two distinct mTORC2-dependent pathways converge on Rac1 to drive breast cancer metastasis. Breast Cancer Res. BCR 19, 74 (2017).

48. Siersbæk, R. et al. IL6/STAT3 Signaling Hijacks Estrogen Receptor α Enhancers to Drive Breast Cancer Metastasis. Cancer Cell 38, 412–423.e9 (2020).

49. Manore, S. G., Doheny, D. L., Wong, G. L. & Lo, H.-W. IL-6/JAK/STAT3 Signaling in Breast Cancer Metastasis: Biology and Treatment. Front. Oncol. 12, 866014 (2022).

50. Ahn, R. et al. The Shc1 adaptor simultaneously balances Stat1 and Stat3 activity to promote breast cancer immune suppression. Nat. Commun. 8, 14638 (2017).

51. Kelly, P. et al. The G12 family of heterotrimeric G proteins promotes breast cancer invasion and metastasis. Proc. Natl. Acad. Sci. U. S. A. 103, 8173–8178 (2006).

52. Rasheed, S. A. K. et al. The emerging roles of Gα12/13 proteins on the hallmarks of cancer in solid tumors. Oncogene 41, 147–158 (2022).

53. Juneja, J. & Casey, P. J. Role of G12 proteins in oncogenesis and metastasis. Br. J. Pharmacol. 158, 32–40 (2009).

54. Gemci, Ö. D. et al. Prognostic Importance of PTEN and P53 in Aggressive Luminal A Subtype Breast Cancers. Eur. J. Breast Health 21, 246–254.

55. Zinn, A. et al. The small GTPase RhoG regulates microtubule-mediated focal adhesion disassembly. Sci. Rep. 9, 5163 (2019).

56. Khongthong, P., Roseweir, A. K. & Edwards, J. The NF-KB pathway and endocrine therapy resistance in breast cancer. Endocr. Relat. Cancer 26, R369–R380 (2019).

57. Yoshihara, K. et al. Inferring tumour purity and stromal and immune cell admixture from expression data. Nat. Commun. 4, 2612 (2013).

58. Timperi, E. et al. Lipid-Associated Macrophages Are Induced by Cancer-Associated Fibroblasts and Mediate Immune Suppression in Breast Cancer. Cancer Res. 82, 3291–3306 (2022).

59. Mehta, A. K., Kadel, S., Townsend, M. G., Oliwa, M. & Guerriero, J. L. Macrophage Biology and Mechanisms of Immune Suppression in Breast Cancer. Front. Immunol. 12, 643771 (2021).

60. Wang, Q. et al. STING agonism reprograms tumor-associated macrophages and overcomes resistance to PARP inhibition in BRCA1-deficient models of breast cancer. Nat. Commun. 13, 3022 (2022).

61. Luca, B. A. et al. Atlas of clinically distinct cell states and ecosystems across human solid tumors. Cell 184, 5482–5496.e28 (2021).

62. Jiang, P. et al. Signatures of T cell dysfunction and exclusion predict cancer immunotherapy response. Nat. Med. 24, 1550–1558 (2018).

63. Jiang, X. et al. The Imprinted Gene PEG3 Inhibits Wnt Signaling and Regulates Glioma Growth. J. Biol. Chem. 285, 8472–8480 (2010).

64. Zhang, M. & Zhang, J. PEG3 mutation is associated with elevated tumor mutation burden and poor prognosis in breast cancer. Biosci. Rep. 40, BSR20201648 (2020).

65. Cingir Koker, S., Jahja, E., Shehwana, H., Keskus, A. G. & Konu, O. Cholinergic Receptor Nicotinic Alpha 5 (CHRNA5) RNAi is associated with cell cycle inhibition, apoptosis, DNA damage response and drug sensitivity in breast cancer. PLoS ONE 13, e0208982 (2018).

66. Zhang, J. et al. Osr2 functions as a biomechanical checkpoint to aggravate CD8+ T cell exhaustion in tumor. Cell 187, 3409–3426.e24 (2024).

67. Tunset, H. M., Feuerherm, A. J., Selvik, L.-K. M., Johansen, B. & Moestue, S. A. Cytosolic Phospholipase A2 Alpha Regulates TLR Signaling and Migration in Metastatic 4T1 Cells. Int. J. Mol. Sci. 20, 4800 (2019).

68. Li, A. et al. Functional diversity of BAI1 (ADGRB1): From angiostasis to synaptic remodeling and disease therapeutics. iScience 29, 114656 (2026).

69. Zhou, J., Wen, W., Xu, Y., Yu, J. & Chen, D. Solute Carrier transporters in tumor metabolism and immune modulation: implications for therapy. J. Transl. Med. 24, 569 (2026).

70. Paul, M. R. et al. Genomic landscape of metastatic breast cancer identifies preferentially dysregulated pathways and targets. J. Clin. Invest. 130, 4252–4265.

71. Hossen, Md. S., Islam, M. S. U., Yasin, M., Ibrahim, M. & Das, A. A Review on the Role of Human Solute Carriers Transporters in Cancer. Health Sci. Rep. 8, e70343 (2025).

72. Ragoussis, J. Regulators of Asymmetric Cell Division in Breast Cancer. Trends Cancer 4, 798–801 (2018).

73. Zamora, I. et al. ONECUT2 is a druggable driver of luminal to basal breast cancer plasticity. Cell. Oncol. 48, 83–99 (2024).

74. Gupta, R., Ponangi, R. & Indresh, K. G. Role of glycosylation in breast cancer progression and metastasis: implications for miRNA, EMT and multidrug resistance. Glycobiology 33, 545–555 (2023).

75. Liu, H. et al. Advances in molecular mechanisms of drugs affecting abnormal glycosylation and metastasis of breast cancer. Pharmacol. Res. 155, 104738 (2020).

76. Durrani, I. A., Bhatti, A. & John, P. The prognostic outcome of ‘type 2 diabetes mellitus and breast cancer’ association pivots on hypoxia-hyperglycemia axis. Cancer Cell Int. 21, 351 (2021).

77. Rose, D. P. & Vona-Davis, L. The cellular and molecular mechanisms by which insulin influences breast cancer risk and progression. Endocr. Relat. Cancer 19, R225–R241 (2012).

78. Javed, S. R., Skolariki, A., Zameer, M. Z. & Lord, S. R. Implications of obesity and insulin resistance for the treatment of oestrogen receptor-positive breast cancer. Br. J. Cancer 131, 1724–1736 (2024).

79. Porta, S. L. et al. Endothelial Tie1–mediated angiogenesis and vascular abnormalization promote tumor progression and metastasis. J. Clin. Invest. 128, 834–845 (2018).

80. Tiainen, L. et al. High baseline Tie1 level predicts poor survival in metastatic breast cancer. BMC Cancer 19, 732 (2019).

81. Chenna, S. S., Gajula, S. N. R. & Nalla, L. V. Polyamine metabolism in cancer: drivers of immune evasion, ferroptosis and therapy resistance. Expert Rev. Mol. Med. 27, e39 (2025).

82. Dustin, D. et al. RON signalling promotes therapeutic resistance in ESR1 mutant breast cancer. Br. J. Cancer 124, 191–206 (2021).

83. Magnussen, S. N. et al. Nephronectin promotes breast cancer brain metastatic colonization via its integrin-binding domains. Sci. Rep. 10, 12237 (2020).

84. Harrington, W. R., Sengupta, S. & Katzenellenbogen, B. S. Estrogen Regulation of the Glucuronidation Enzyme UGT2B15 in Estrogen Receptor-Positive Breast Cancer Cells. Endocrinology 147, 3843–3850 (2006).

85. Luo, L. et al. Distinct lymphocyte antigens 6 (Ly6) family members Ly6D, Ly6E, Ly6K and Ly6H drive tumorigenesis and clinical outcome. Oncotarget 7, 11165–11193 (2016).

86. Pratt, S. J., Hernández-Ochoa, E. & Martin, S. S. Calcium signaling: breast cancer’s approach to manipulation of cellular circuitry. Biophys. Rev. 12, 1343–1359 (2020).

87. Wang, Y. et al. TRPC3-mediated NFATc1 calcium signaling promotes triple negative breast cancer migration through regulating glypican-6 and focal adhesion. Pflugers Arch. 477, 253–272 (2025).

88. Su, S. et al. A Positive Feedback Loop between Mesenchymal-like Cancer Cells and Macrophages Is Essential to Breast Cancer Metastasis. Cancer Cell 25, 605–620 (2014).

89. Guo, Y. et al. Zeb1 induces immune checkpoints to form an immunosuppressive envelope around invading cancer cells. Sci. Adv. 7, eabd7455 (2021).

90. Mariathasan, S. et al. TGFβ attenuates tumour response to PD-L1 blockade by contributing to exclusion of T cells. Nature 554, 544–548 (2018).

91. Liu, D., Vadgama, J. & Wu, Y. Basal-like breast cancer with low TGFβ and high TNFα pathway activity is rich in activated memory CD4 T cells and has a good prognosis. Int. J. Biol. Sci. 17, 670–682 (2021).

92. Drasin, D. J., Robin, T. P. & Ford, H. L. Breast cancer epithelial-to-mesenchymal transition: examining the functional consequences of plasticity. Breast Cancer Res. 13, 226 (2011).

93. Drabsch, Y. & ten Dijke, P. TGF-β Signaling in Breast Cancer Cell Invasion and Bone Metastasis. J. Mammary Gland Biol. Neoplasia 16, 97–108 (2011).

94. Ye, J. et al. Fibroblast Growth Factor Receptor 4 Promotes Triple-Negative Breast Cancer Progression via Regulating Fatty Acid Metabolism Through the AKT/RYR2 Signaling. Cancer Med. 13, e70439 (2024).

95. Shin, D. S. et al. Primary resistance to PD-1 blockade mediated by JAK1/2 mutations. Cancer Discov. 7, 188–201 (2017).

96. Hammerl, D. et al. Spatial immunophenotypes predict response to anti-PD1 treatment and capture distinct paths of T cell evasion in triple negative breast cancer. Nat. Commun. 12, 5668 (2021).

97. Okano, M. et al. Triple-Negative Breast Cancer with High Levels of Annexin A1 Expression Is Associated with Mast Cell Infiltration, Inflammation, and Angiogenesis. Int. J. Mol. Sci. 20, 4197 (2019).

98. Maltby, S., Khazaie, K. & McNagny, K. M. Mast Cells in Tumor Growth: Angiogenesis, Tissue Remodeling and Immune-modulation. Biochim. Biophys. Acta 1796, 19–26 (2009).

99. Glajcar, A. et al. The relationship between breast cancer molecular subtypes and mast cell populations in tumor microenvironment. Virchows Arch. 470, 505–515 (2017).

100. Sousa, S. et al. Human breast cancer cells educate macrophages toward the M2 activation status. Breast Cancer Res. BCR 17, 101 (2015).

101. Zwager, M. C. et al. Assessing the role of tumour-associated macrophage subsets in breast cancer subtypes using digital image analysis. Breast Cancer Res. Treat. 198, 11–22 (2023).

102. Wang, S. et al. Targeting M2-like tumor-associated macrophages is a potential therapeutic approach to overcome antitumor drug resistance. Npj Precis. Oncol. 8, 31 (2024).

103. Hachim, M. Y., Hachim, I. Y., Talaat, I. M., Yakout, N. M. & Hamoudi, R. M1 Polarization Markers Are Upregulated in Basal-Like Breast Cancer Molecular Subtype and Associated With Favorable Patient Outcome. Front. Immunol. 11, 560074 (2020).

104. Hachim, M. Y., Talaat, I. & Hachim, I. Y. Macrophage M2 polarization markers are downregulated in Basal compared to Luminal A and Luminal B Breast Cancer. FASEB J. 34, 1–1 (2020).

105. Oshi, M. et al. Intra-Tumoral Angiogenesis Is Associated with Inflammation, Immune Reaction and Metastatic Recurrence in Breast Cancer. Int. J. Mol. Sci. 21, 6708 (2020).

106. Luca, B. A. et al. Atlas of clinically distinct cell states and ecosystems across human solid tumors. Cell 184, 5482–5496.e28 (2021).

107. Troester, M. A. et al. Activation of Host Wound Responses in Breast Cancer Microenvironment. Clin. Cancer Res. 15, 7020–7028 (2009).

108. Stewart, D. A., Yang, Y., Makowski, L. & Troester, M. A. Basal-like Breast Cancer Cells Induce Phenotypic and Genomic Changes in Macrophages. Mol. Cancer Res. 10, 727–738 (2012).

109. Arnold, K. M., Opdenaker, L. M., Flynn, D. & Sims-Mourtada, J. Wound Healing and Cancer Stem Cells: Inflammation as a Driver of Treatment Resistance in Breast Cancer. Cancer Growth Metastasis 8, 1–13 (2015).

110. Loi, S. et al. Tumor-Infiltrating Lymphocytes and Prognosis: A Pooled Individual Patient Analysis of Early-Stage Triple-Negative Breast Cancers. J. Clin. Oncol. 37, 559–569 (2019).

111. Adams, S. et al. Prognostic value of tumor-infiltrating lymphocytes in triple-negative breast cancers from two phase III randomized adjuvant breast cancer trials: ECOG 2197 and ECOG 1199. J. Clin. Oncol. Off. J. Am. Soc. Clin. Oncol. 32, 2959–2966 (2014).

112. Gruosso, T. et al. Spatially distinct tumor immune microenvironments stratify triple-negative breast cancers. J. Clin. Invest. 129, 1785–1800 (2019).

113. Oshi, M. et al. Enhanced immune response outperform aggressive cancer biology and is associated with better survival in triple-negative breast cancer. Npj Breast Cancer 8, 1–10 (2022).

114. Oshi, M. et al. Immune cytolytic activity is associated with reduced intra-tumoral genetic heterogeneity and with better clinical outcomes in triple negative breast cancer. Am. J. Cancer Res. 11, 3628–3644 (2021).

115. Yam, C. et al. Immune Phenotype and Response to Neoadjuvant Therapy in Triple-Negative Breast Cancer. Clin. Cancer Res. 27, 5365–5375 (2021).

116. Mao, Y. et al. The Prognostic Value of Tumor-Infiltrating Lymphocytes in Breast Cancer: A Systematic Review and Meta-Analysis. PLOS ONE 11, e0152500 (2016).

117. Chung, Y. R., Kim, H. J., Jang, M. H. & Park, S. Y. Prognostic value of tumor infiltrating lymphocyte subsets in breast cancer depends on hormone receptor status. Breast Cancer Res. Treat. 161, 409–420 (2017).

118. Egelston, C. A. et al. Tumor-infiltrating exhausted CD8+ T cells dictate reduced survival in premenopausal estrogen receptor–positive breast cancer. JCI Insight 7, e153963.

119. Castellaro, A. M., Rodriguez-Baili, M. C., Di Tada, C. E. & Gil, G. A. Tumor-Associated Macrophages Induce Endocrine Therapy Resistance in ER+ Breast Cancer Cells. Cancers 11, 189 (2019).

120. Tharp, K. M. et al. Tumor-associated macrophages restrict CD8+ T cell function through collagen deposition and metabolic reprogramming of the breast cancer microenvironment. Nat. Cancer 5, 1045–1062 (2024).

121. Munir, M. T. et al. Tumor-Associated Macrophages as Multifaceted Regulators of Breast Tumor Growth. Int. J. Mol. Sci. 22, 6526 (2021).

122. Pellegrino, B. et al. Luminal Breast Cancer: Risk of Recurrence and Tumor-Associated Immune Suppression. Mol. Diagn. Ther. 25, 409–424 (2021).

123. Aouad, P. et al. Epithelial-mesenchymal plasticity determines estrogen receptor positive breast cancer dormancy and epithelial reconversion drives recurrence. Nat. Commun. 13, 4975 (2022).

124. El-Botty, R. et al. Oxidative phosphorylation is a metabolic vulnerability of endocrine therapy and palbociclib resistant metastatic breast cancers. Nat. Commun. 14, 4221 (2023).

125. Keoh, L. Q., Chiu, C.-F. & Ramasamy, T. S. Metabolic Plasticity and Cancer Stem Cell Metabolism: Exploring the Glycolysis-OXPHOS Switch as a Mechanism for Resistance and Tumorigenesis. Stem Cell Rev. Rep. 21, 2446–2468 (2025).

126. Ginestier, C. et al. Retinoid signaling regulates breast cancer stem cell differentiation. Cell Cycle Georget. Tex 8, 3297–3302 (2009).

127. BeLow, M. & Osipo, C. Notch Signaling in Breast Cancer: A Role in Drug Resistance. Cells 9, 2204 (2020).

128. Kasper, M., Jaks, V., Fiaschi, M. & Toftgård, R. Hedgehog signalling in breast cancer. Carcinogenesis 30, 903–911 (2009).

129. Powell, E., Piwnica-Worms, D. & Piwnica-Worms, H. Contribution of p53 to Metastasis. Cancer Discov. 4, 405–414 (2014).

130. Marvalim, C., Datta, A. & Lee, S. C. Role of p53 in breast cancer progression: An insight into p53 targeted therapy. Theranostics 13, 1421–1442 (2023).

131. Bertheau, P. et al. p53 in breast cancer subtypes and new insights into response to chemotherapy. The Breast 22, S27–S29 (2013).

132. Mueller, S. et al. p53 Expression in Luminal Breast Cancer Correlates With *TP53* Mutation and Primary Endocrine Resistance. Mod. Pathol. 36, 100100 (2023).

133. Berger, C. E., Qian, Y., Liu, G., Chen, H. & Chen, X. p53, a Target of Estrogen Receptor (ER) α, Modulates DNA Damage-induced Growth Suppression in ER-positive Breast Cancer Cells*. J. Biol. Chem. 287, 30117–30127 (2012).

134. Nunnery, S. E. & Mayer, I. A. Targeting the PI3K/AKT/mTOR Pathway in Hormone Positive Breast Cancer. Drugs 80, 1685–1697 (2020).

135. Rudolph, M. et al. A hedgehog pathway-dependent gene signature is associated with poor clinical outcomes in Luminal A breast cancer. Breast Cancer Res. Treat. 169, 457–467 (2018).

136. Adibfar, S. et al. The molecular mechanisms and therapeutic potential of EZH2 in breast cancer. Life Sci. 286, 120047 (2021).

137. Oshi, M. et al. G2M Cell Cycle Pathway Score as a Prognostic Biomarker of Metastasis in Estrogen Receptor (ER)-Positive Breast Cancer. Int. J. Mol. Sci. 21, 2921 (2020).

138. Cheang, M. C. U. et al. Ki67 Index, HER2 Status, and Prognosis of Patients With Luminal B Breast Cancer. JNCI J. Natl. Cancer Inst. 101, 736–750 (2009).

139. Yang, J. et al. Estrogen receptor-α directly regulates the hypoxia-inducible factor 1 pathway associated with antiestrogen response in breast cancer. Proc. Natl. Acad. Sci. U. S. A. 112, 15172–15177 (2015).

140. Goncalves, A., Finetti, P., Birnbaum, D. & Bertucci, F. The CINSARC signature predicts the clinical outcome in patients with Luminal B breast cancer. Npj Breast Cancer 7, 48 (2021).

141. Borg, A., Zhang, Q. X., Olsson, H. & Wenngren, E. Chromosome 1 alterations in breast cancer: allelic loss on 1p and 1q is related to lymphogenic metastases and poor prognosis. Genes. Chromosomes Cancer 5, 311–320 (1992).

142. Goh, J. Y. et al. Chromosome 1q21.3 amplification is a trackable biomarker and actionable target for breast cancer recurrence. Nat. Med. 23, 1319–1330 (2017).

143. Liu, Y. et al. Genomic Copy Number Imbalances Associated with Bone and Non-bone Metastasis of Early-Stage Breast Cancer. Breast Cancer Res. Treat. 143, 189–201 (2014).

144. Muthuswami, M. et al. Breast Tumors with Elevated Expression of 1q Candidate Genes Confer Poor Clinical Outcome and Sensitivity to Ras/PI3K Inhibition. PLoS ONE 8, e77553 (2013).

145. Orsetti, B. et al. Genetic profiling of chromosome 1 in breast cancer: mapping of regions of gains and losses and identification of candidate genes on 1q. Br. J. Cancer 95, 1439–1447 (2006).

146. Gerashchenko, T. S. et al. The Activity of KIF14, Mieap, and EZR in a New Type of the Invasive Component, Torpedo-Like Structures, Predetermines the Metastatic Potential of Breast Cancer. Cancers 12, 1909 (2020).

147. Corson, T. W. & Gallie, B. L. KIF14 mRNA expression is a predictor of grade and outcome in breast cancer. Int. J. Cancer 119, 1088–1094 (2006).

148. Nelson, D. J. et al. A review of the importance of immune responses in luminal B breast cancer. Oncoimmunology 6, e1282590 (2017).

149. Moura, T. et al. Early-Stage Luminal B-like Breast Cancer Exhibits a More Immunosuppressive Tumor Microenvironment than Luminal A-like Breast Cancer. Biomolecules 15, (2025).

150. Jiang, Y. et al. Comparing efficacy of neoadjuvant therapy of triple-negative breast cancer: A Bayesian network meta-regression analysis. Medicine (Baltimore) 105, e46962 (2026).

151. Martín, M. et al. TNBC-DX genomic test in early-stage triple-negative breast cancer treated with neoadjuvant taxane-based therapy☆. Ann. Oncol. 36, 158–171 (2025).

152. Pan, H. et al. 20-Year Risks of Breast-Cancer Recurrence after Stopping Endocrine Therapy at 5 Years. N. Engl. J. Med. 377, 1836–1846 (2017).

153. Strasser-Weippl, K., Badovinac-Crnjevic, T., Fan, L. & Goss, P. E. Extended adjuvant endocrine therapy in hormone-receptor positive breast cancer. The Breast 22, S171–S175 (2013).

154. Parker, J. S. et al. Supervised Risk Predictor of Breast Cancer Based on Intrinsic Subtypes. J. Clin. Oncol. 27, 1160–1167 (2009).

155. Chen, L. et al. Body mass index and risk of luminal, HER2-overexpressing, and triple negative breast cancer. Breast Cancer Res. Treat. 157, 545–554 (2016).

156. Chen, L. et al. Reproductive factors and risk of luminal, HER2-overexpressing and triple negative breast cancer among multiethnic women. Cancer Epidemiol. Biomark. Prev. Publ. Am. Assoc. Cancer Res. Cosponsored Am. Soc. Prev. Oncol. 25, 1297–1304 (2016).

157. Hamilton, A. M. et al. The landscape of immune microenvironments in racially-diverse breast cancer patients. Cancer Epidemiol. Biomark. Prev. Publ. Am. Assoc. Cancer Res. Cosponsored Am. Soc. Prev. Oncol. 31, 1341–1350 (2022).

158. Wong, D. J. et al. Module Map of Stem Cell Genes Guides Creation of Epithelial Cancer Stem Cells. Cell Stem Cell 2, 333–344 (2008).

159. Liu, R. et al. The Prognostic Role of a Gene Signature from Tumorigenic Breast-Cancer Cells. N. Engl. J. Med. 356, 217–226 (2007).

160. Goncalves, A., Finetti, P., Birnbaum, D. & Bertucci, F. The CINSARC signature predicts the clinical outcome in patients with Luminal B breast cancer. Npj Breast Cancer 7, 48 (2021).

161. Creighton, C. J. et al. Genes regulated by estrogen in breast tumor cells in vitro are similarly regulated in vivoin tumor xenografts and human breast tumors. Genome Biol. 7, R28 (2006).

162. Zhang, B. et al. An EMT-Related Gene Signature to Predict the Prognosis of Triple-Negative Breast Cancer. Adv. Ther. 40, 4339–4357 (2023).

163. Cui, Y. et al. A novel epithelial-mesenchymal transition (EMT)-related gene signature of predictive value for the survival outcomes in lung adenocarcinoma. Front. Oncol. 12, 974614 (2022).

164. Newman, A. M. et al. Robust enumeration of cell subsets from tissue expression profiles. Nat. Methods 12, 453–457 (2015).

165. Diossy, M. et al. Strand Orientation Bias Detector to determine the probability of FFPE sequencing artifacts. Brief. Bioinform. 22, bbab186 (2021).

166. Ullah, I. et al. Evolutionary history of metastatic breast cancer reveals minimal seeding from axillary lymph nodes. J. Clin. Invest. 128, 1355–1370.

167. Li, X. et al. PathwayTMB: A pathway-based tumor mutational burden analysis method for predicting the clinical outcome of cancer immunotherapy. Mol. Ther. Nucleic Acids 34, 102026 (2023).

